# Repeatability and timing of tropical influenza epidemics

**DOI:** 10.1101/2022.11.04.22281944

**Authors:** Joseph L Servadio, Pham Quang Thai, Marc Choisy, Maciej F Boni

**Affiliations:** Center for Infectious Disease Dynamics and Department of Biology, Pennsylvania State University, University Park, PA, USA; National Institute of Hygiene and Epidemiology, Hanoi, Vietnam; School of Preventative Medicine and Public Health, Hanoi Medical University, Hanoi, Vietnam; Oxford University Clinical Research Unit, Ho Chi Minh City, Vietnam; Centre for Tropical Medicine and Global Health, Nuffield Department of Medicine, University of Oxford, Oxford, UK

**Keywords:** influenza, seasonality, repeatability, Vietnam, SIRS model

## Abstract

Much of the world experiences influenza in yearly recurring seasons, particularly in temperate areas. These patterns can be considered repeatable, occurring predictably and consistently. In tropical areas, including southeast Asia, this consistency is less conspicuous. This study aimed to assess repeatability of influenza in Vietnam. A mathematical model was developed incorporating periods of increased transmission, and fit to data from sentinel hospitals throughout Vietnam as well as four temperate locations. Repeatability was evaluated through the variance of the timings of peak transmission. Model fits from Vietnam show high variance (sd = 70-171) in peak transmission timing; peaks occurred at irregular intervals and throughout different times of year. Fits from temperate locations showed regular, annual epidemics in winter months, with low variance in peak timings (sd = 33-74). This suggests that influenza patterns are not repeatable or seasonal in Vietnam. Influenza prevention in Vietnam therefore cannot rely anticipating regularly occurring outbreaks.

## 1. Introduction

Influenza is consistently regarded as a major public health concern globally, causing high burden despite efforts to update and distribute vaccines prior to upcoming influenza seasons (1). It has been estimated that, annually, between 10 and 20 percent of the global population is infected with influenza (2), and between 250,000 and 650,000 deaths (3–5) occur. Morbidity and mortality differ across age groups, with young children and elderly adults experiencing highest burden (6,7). In addition to morbidity and mortality, influenza has been shown to adversely impact the global economy (8). The public health importance of influenza is apparent both during typical influenza seasons as well as during major pandemics, as seen notably in 1918 as well as in 1957, 1968, and most recently in 2009 (9).

In much of the world, influenza epidemics are viewed as regularly occurring disease events, with temperate regions of the world reporting increases in influenza incidence during winter months (10,11). Several studies based in North America and Europe incorporate annual seasonality of influenza as a known component of influenza dynamics (12–14). This is a well-regarded pattern outside of influenza as well; other respiratory diseases with identified annual or near-annual patterns in temperate regions include RSV (15), common cold causing coronaviruses (16,17), and rhinoviruses (18).

For any pathogen, if periodic increases in incidence occur and align with the calendar year, a natural conclusion to draw is that the disease dynamics are influenced by annually recurring external factors. When periodic signals are observed in disease transmission, as is seen with influenza in temperate regions of the world, it is common to reinforce the notion of seasonality by using periodic events and phenomena to describe the dynamics of the disease, commonly through statistical or mathematical models. Within influenza or other respiratory viruses, this has been seen by using annual cyclic predictors such as climate patterns (7,19), school terms (20), and holidays (21) to predict disease incidence. In regions that experience cyclic patterns in weather, seasonal changes in human behavior typically follow. Seasonal behavioral changes can include some that affect influenza transmission, such as gathering in indoor spaces more frequently. Additionally, virus transmission in different climate settings, such as under different humidity and temperature conditions, have been examined (22,23). While both of these potential mechanisms of transmission, social contact patterns and virus survival in the environment, have been studied previously, there currently is no consensus of which mechanism or set of mechanisms has the strongest influence on seasonal influenza patterns (24). There is continuing debate whether observed seasonality relates more strongly to seasonal changes in weather patterns or to seasonal changes in human behavior (25,26).

In contrast, tropical regions of the world do not experience pronounced changes in temperature patterns and also do not appear to experience strong influenza seasonality, though a consensus has yet to be reached on this topic. In much of the tropics, influenza is seen throughout the course of the year rather than during particular parts of the year (1,7,27–29). Less consistent evidence of influenza seasonality has been observed in tropical countries, including much of Latin America (30), sub-Saharan Africa (31), and south and southeast Asia (32–34). Some work has been done seeking to determine whether influenza follows annual or nonannual cyclic patterns; results of these studies have provided some evidence for annual cycles or nonannual cycles (35–38), while others have not found substantial evidence for cyclic patterns in influenza (39–41).

Despite less consistent seasonality in the tropics, many studies have aimed to relate influenza patterns to climate in the tropics. Some evidence has shown that environmental factors may influence influenza in both a tropical and temperate setting, but in different ways (28,42). Differences in seasonal changes in precipitation and humidity as well as seasonal changes in behaviors such as social mixing may relate to the differences in distinctness of influenza seasons. Despite ample literature showing statistical associations between influenza (or other respiratory viruses) and seasonal covariates, notably temperature, precipitation, humidity, and wind speed (35,43–47), there is a dearth of research seeking to assess quantitative support for the existence of seasonal or annual patterns in the tropics (48).

In describing seasonality of influenza and, as a result, influenza seasons, definitions of seasonality can differ across studies. While many studies refer to seasonality as regularly occurring events during a particular time of year, others view seasonality as an annually recurring event, even if the time of year is not consistent. An important difference is that inconsistent seasonality, such as observing an outbreak every year, but at a different time each year, does not describe a system that is predictable through repetition. Here, we define “repeatability” as seasonality that occurs at a consistent time of year. In a repeatable system, knowledge of one season occurring is highly informative of when the next season will occur. Repeatability of influenza refers to having consistent and predictable seasonality at the same time each year.

This study aimed to determine the presence or absence of annual seasonality, or repeatability, of influenza in the tropics, using ten years of standardized surveillance data from Vietnam as a case study. Our starting hypothesis is that influenza is seasonal in Vietnam. We developed a mathematical model that describes influenza dynamics in Vietnam and incorporates parameters representing periods of increased transmission. Our statistical framework includes parameters representing peak timings of transmission and the repeatability of these peak timings is evaluated through a Bayesian inference. Fitting the model to weekly influenza time series from four temperate locations (Netherlands, Denmark, and two regions of the United States) as well as northern, central, and southern Vietnam allows us to assess whether periods of increased transmission occur with greater or lesser repeatability and compare between temperate and tropical locations. The results of this study provide insight into whether influenza dynamics in Vietnam exhibit any annual or nonannual repeatable cycles.

## 2. Results

Fifteen hospitals throughout northern, central, and southern Vietnam provided data used for this study (Fig 1). Between 2006 and 2015, 4,799,182 outpatients attended the fifteen hospitals participating in Vietnam’s National Influenza Sentinel Surveillance (1,644,488 in northern VN, 908,057 in central VN, 2,246,637 in southern VN), 515,598 of whom were ILI patients (214,238 in northern VN, 92,155 in central VN, 209,205 in southern VN). A total of 43,878 molecular-diagnostic tests were administered (18,190 in northern VN, 12,631 in central VN, and 13,057 in southern VN), of which 9,204 (21.0%) were PCR-positive for influenza virus. In northern Vietnam, 3,599 (19.8%) of tests were flu-positive, with 13.1% positive for influenza A, 5.8% positive for A/H1, 7.3% positive for A/H3, and 6.7% positive for influenza B. In central Vietnam, 2,634 (20.9%) of tests were positive, with 13.5% positive for influenza A, 6.9% positive for A/H1, 6.6% positive for A/H3, and 7.3% positive for influenza B. In southern Vietnam, 2,971 (22.8%) of tests were positive, with 14.8% positive for influenza A, 6.9% positive for A/H1, 7.9% positive for A/H3, and 8.0% positive for influenza B. A total of 22 tests were positive for both influenza A and influenza B, and eight tests were positive for influenza A without specification of subtype. These data points were discarded. The detrended time series of ILI+ stratified by (sub)type are shown in Figure 2.

**Figure 1.**
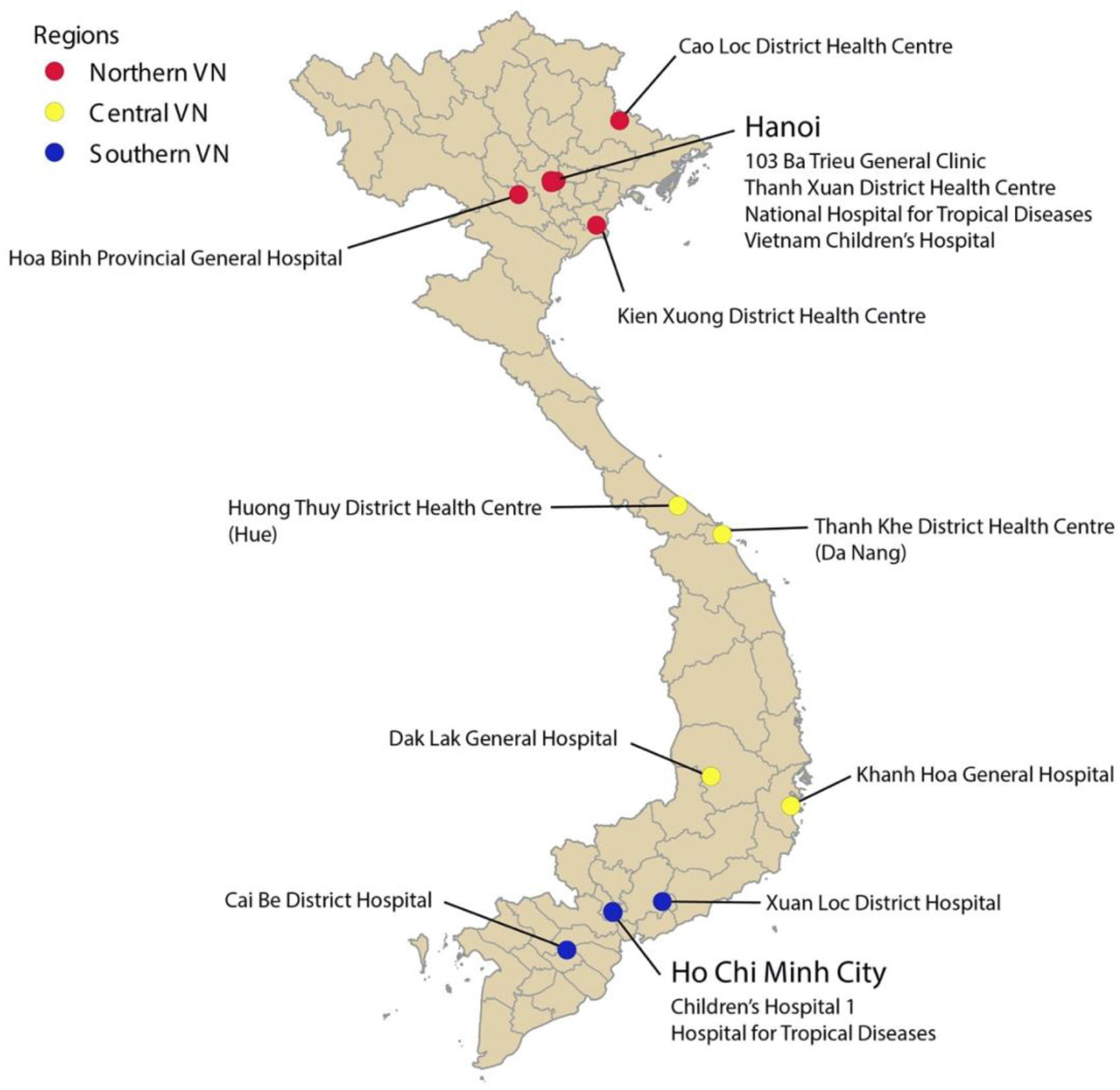
Locations of sentinel hospitals in Vietnam. Sites are colored by region: northern, central, and southern Vietnam. Sites located in the cities of Hanoi and Ho Chi Minh City are listed underneath the city name.

**Figure 2.**
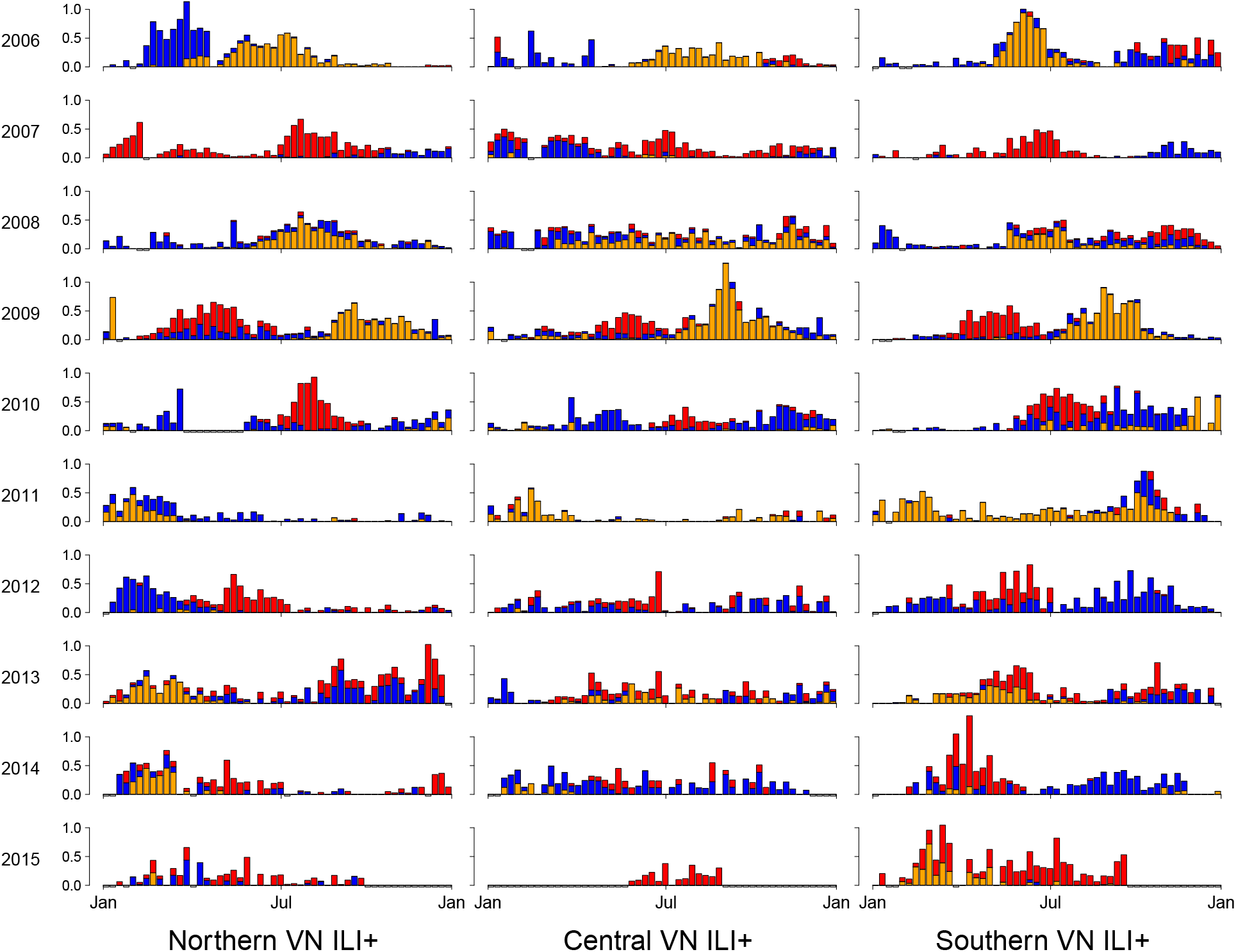
Stacked bar chart of weekly time series of detrended ILI+. Detrended ILI+ is shown for each year between 2006 (top row) and 2015 (bottom row) for each of northern (left column), central (middle column), and southern (right column) Vietnam. Colors denote (sub)types of influenza, with orange representing A/H1, red representing A/H3, and blue representing B.

### 2.1. Descriptive indicators of seasonality

Autocorrelation functions showed inconsistent and weak evidence of annual or nonannual cycles in the influenza data from Vietnam for all (sub)types as well as for combined ILI+. In contrast, the four temperate locations showed strong positive correlations for lags at and near one year (Figure S1). Similarly, the Morlet wavelets showed inconsistency in detecting repeated cycles in the data from Vietnam. While some periodicity was detected, the duration of periods differed across locations and (sub)types, and no periodicity was detected that consistently spanned the entire time period. The wavelets for the temperate locations, however, showed consistently strong evidence of seasonality, with 52-week cycles providing a strong fit over the entire time span of all four locations (Figure S2). These two descriptive analyses provide preliminary evidence that seasonality of influenza is much weaker in Vietnam compared to temperate locations.

### 2.2. Influenza peak seasonality and repeatability in Vietnam

The model fits showed close fits for the individual types and subtypes of influenza across the three regions of Vietnam. The fit to combined ILI+ was less accurate in comparison. Less precision was observed in the overall fit for influenza type B, and the fits for the individual A subtypes most closely matched the observed data (Figure 3).

**Figure 3.**
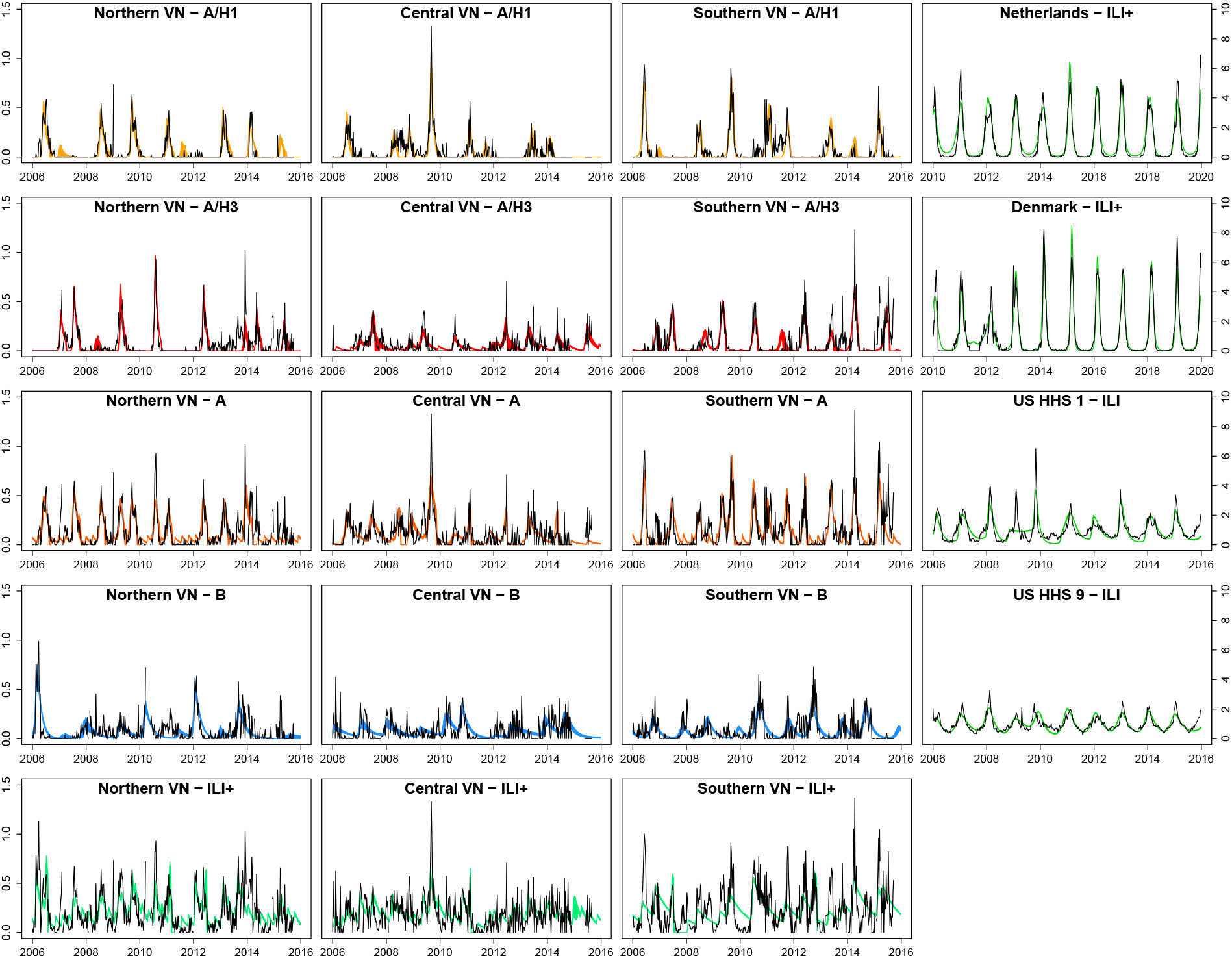
Posterior model fits across influenza (sub)types as well as for combined ILI+. Columns from left to right show model fits for northern Vietnam, central Vietnam, southern Vietnam, and four temperate locations. The colored bands show all fitted trajectories from the posterior parameter samples, with different (sub)types represented by different colors, and black lines show the observed data.

The estimates of the *τ*_*k*_ parameters, representing spacings between epidemic peaks, in Vietnam varied across locations and among subtypes. The average space between peaks ranged between 246 days (ILI+ in central VN) and 399 days (A/H1 in northern VN). The standard deviations of the time between peaks ranged between 70 days (B in southern VN) and 171 days (A/H1 in central VN) (Table 1, Figure 4). These high standard deviations in Vietnam indicate generally weak seasonality in transmission.

**Table 1.**
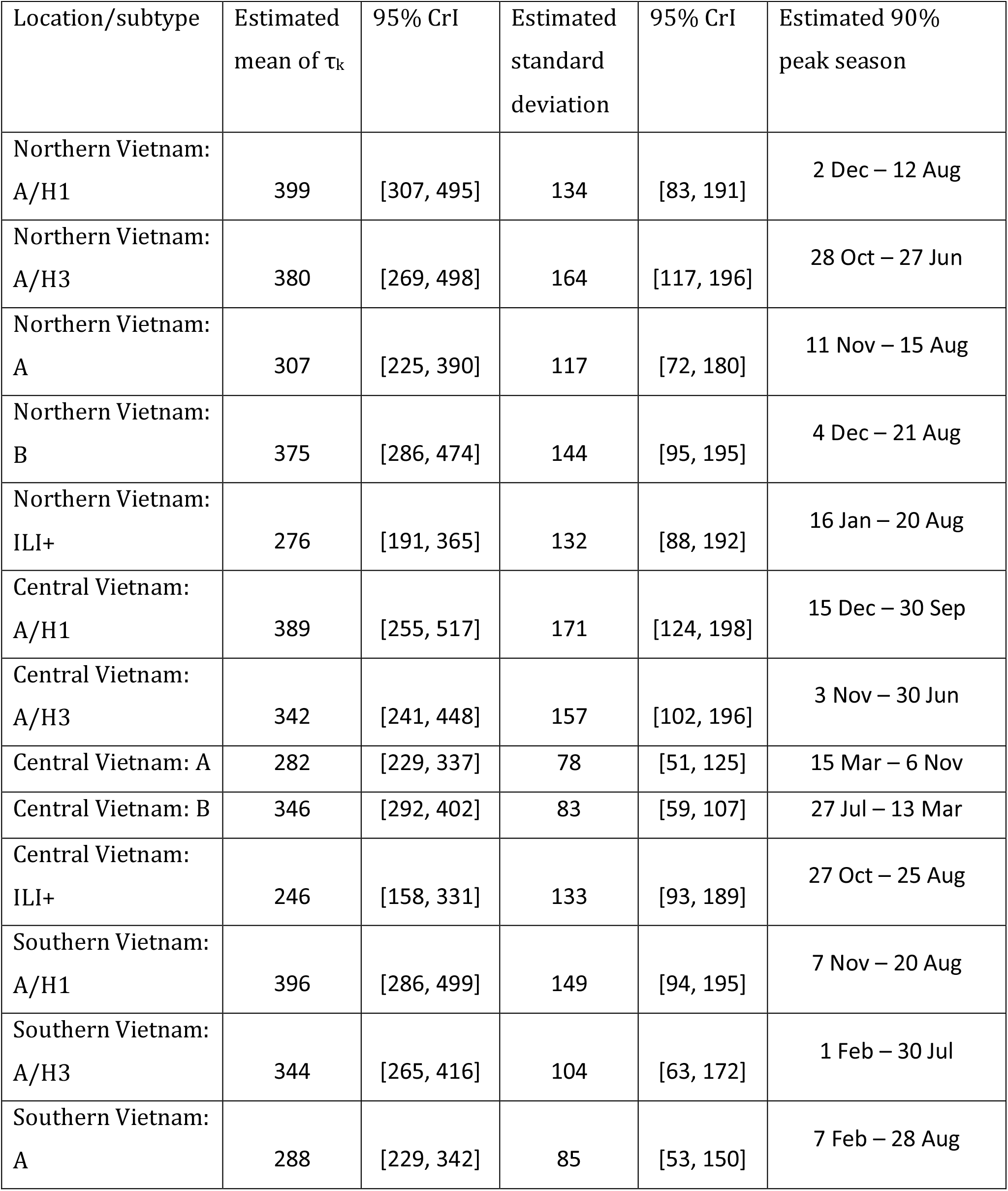
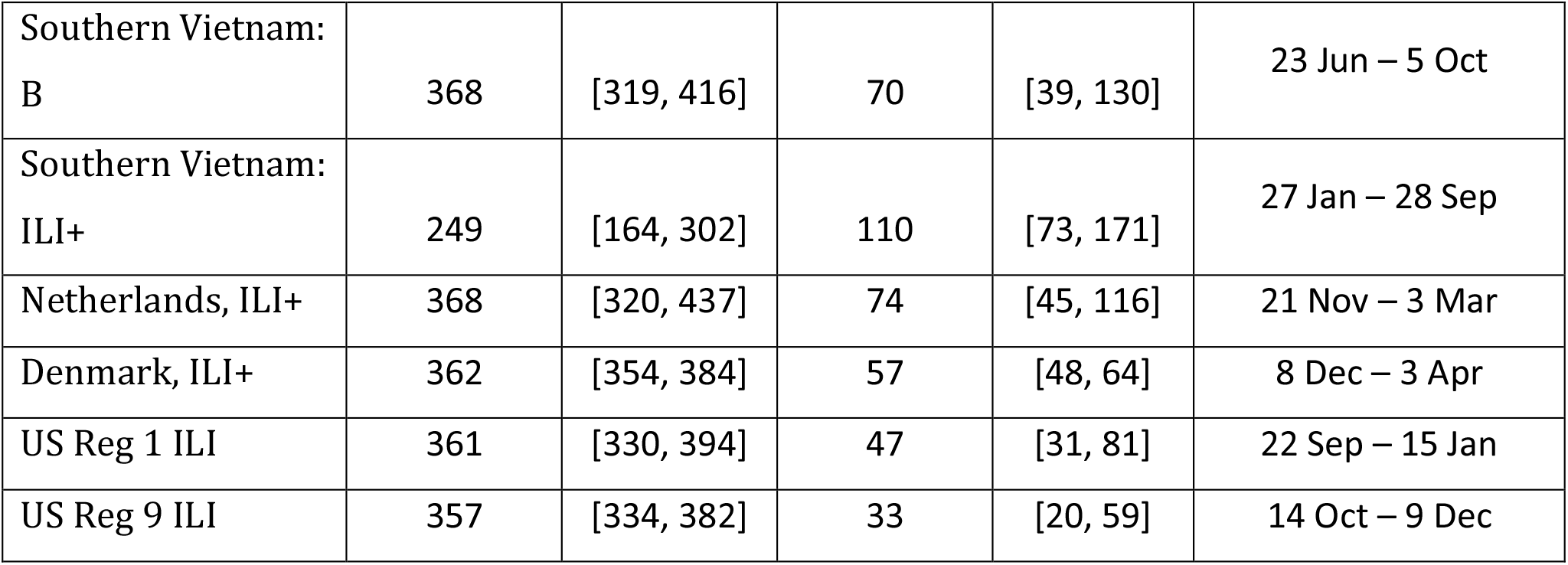
Estimated mean and standard deviation of spacings between transmission peaks with 95% credible intervals. Estimated peak seasons represent the shortest time window that includes 90% of the posterior density of the *φ*_*k*_.

**Figure 4.**
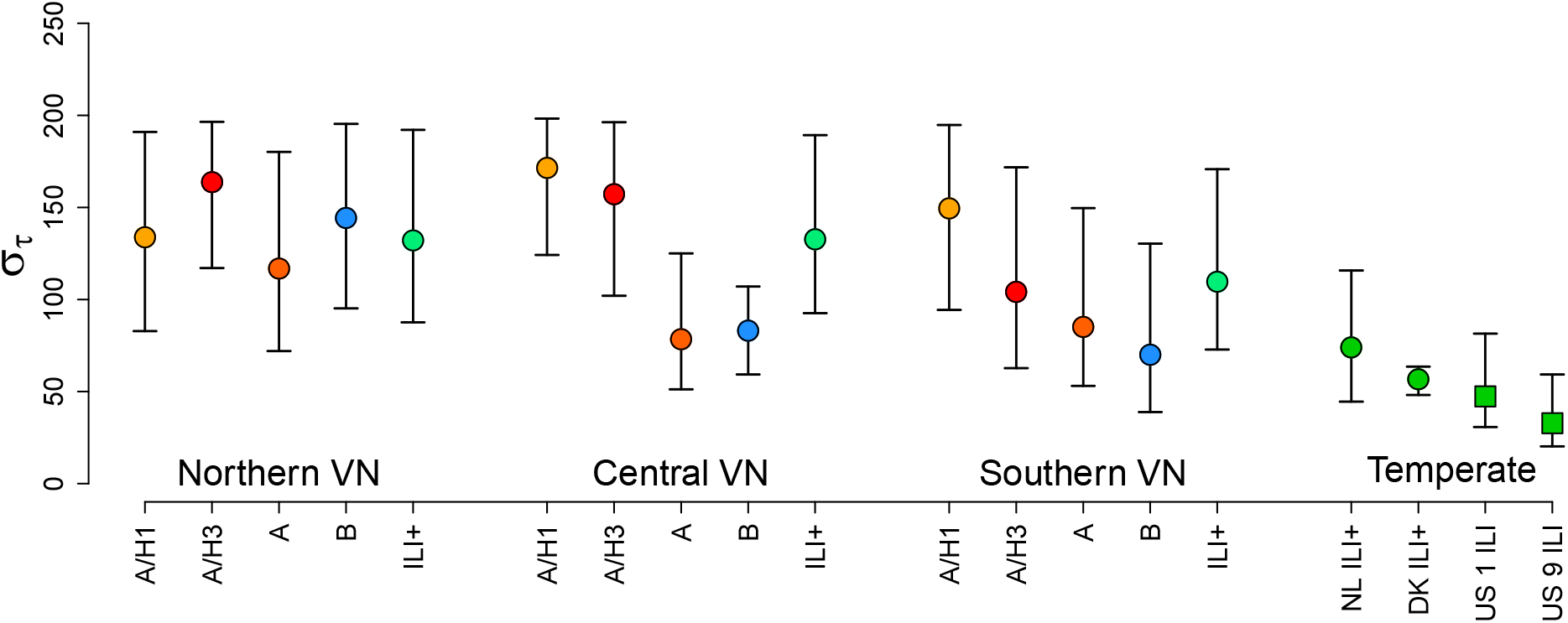
Estimated standard deviations for peak season timing (φ_k_)(medians from posterior distributions, with 95% credible intervals represented by error bars) for A/H1, A/H3, combined A, B, and combined ILI+ in northern, central, and southern Vietnam. These are compared to estimated standard deviations for timings of ILI+ peaks in the Netherlands and Denmark and ILI in two regions of the United States.

Converting the estimated *τ*_*k*_ parameters into *φ*_*k*_ parameters (representing timings of peaks) shows whether certain times of year are most common for influenza peaks. These *φ*_*k*_ parameters can be converted into calendar dates, and times of year when the *φ*_*k*_ most frequently occur would represent a regular “peak season” of influenza, if consistent. If influenza is repeatable, then the timings of peaks would be consistent, and the peak season would cover a short span of time. Across the three regions of Vietnam, peaks are inconsistent, occurring throughout the course of the calendar year (Figure 5). Estimated peak seasons in Table 1 and Figure 5 represent the minimum time window that would encompass 90% of the posterior distribution of all *φ*_*k*_. Within Vietnam, the shortest of these windows was 104 days, ranging between 23 Jun – 5 Oct for influenza B in southern Vietnam. The longest window was 302 days, ranging between 27 Oct – 25 Aug for ILI+ in central Vietnam. The median window was 242 days.

**Figure 5.**
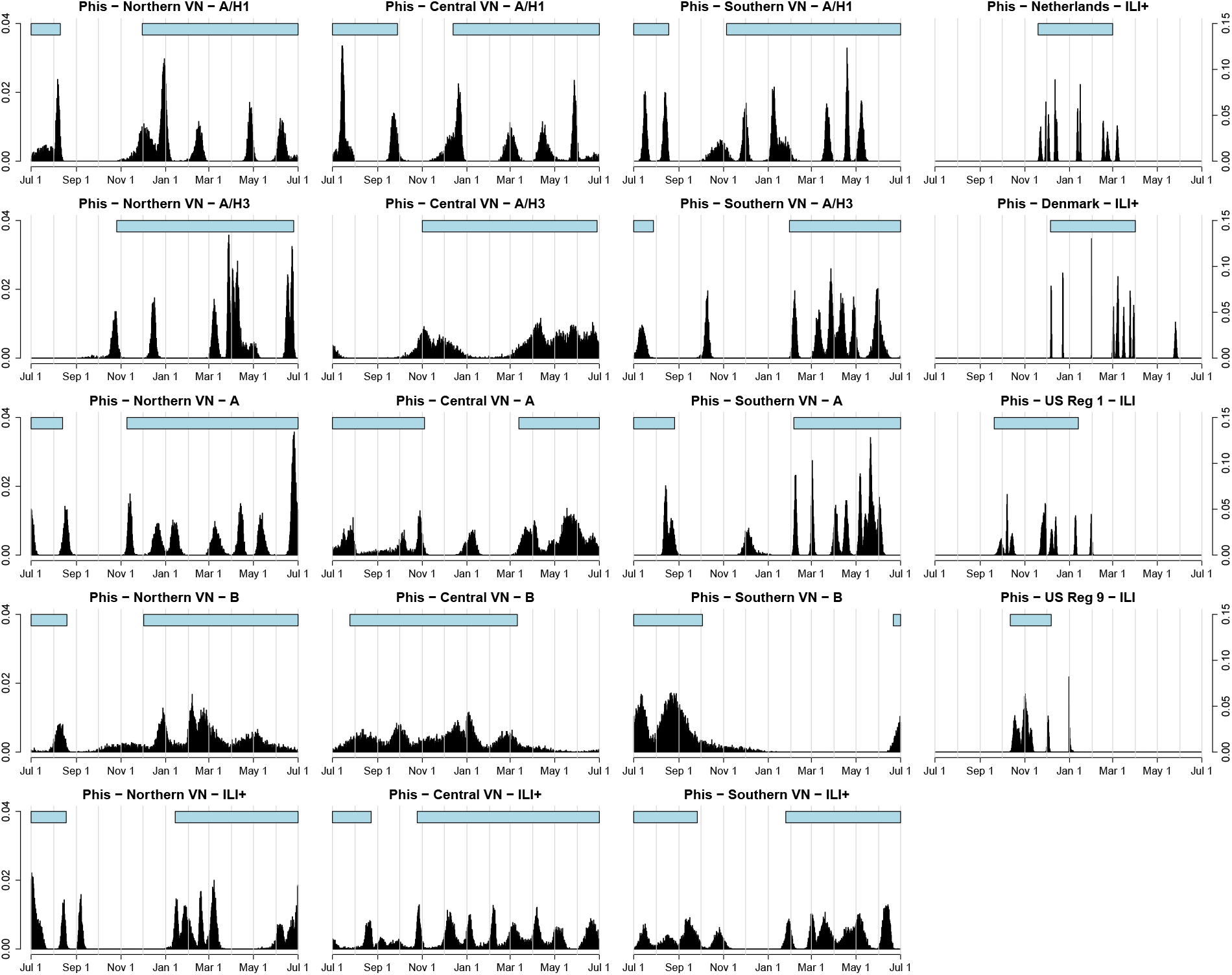
Posterior densities for timing of peak influenza transmission, i.e. the posterior distributions of *φ*_*k*_ – the peak time – across all seasons included in the analysis. Density is determined by frequency of *φ*_*k*_ values in posterior MCMC samples corresponding to days of the calendar year, spanning between July 1 and June 30. Vertical lines show first days of calendar months. Horizontal bars above the distribution show the minimum time span that encompasses at least 90% of the posterior density, serving as a representation for a peak season.

### 2.3. Influenza peak seasonality and repeatability in temperate countries

The model shows close fits to the data from the four temperate locations (Fig 3). The time elapsing between transmission peaks was more consistent among temperate regions compared to Vietnam. The average time elapsed was 368 days (95% CrI: [320, 437]) for the Netherlands, 362 days (95% CrI: [354, 384]) for Denmark, 361 days (95% CrI: [330, 394]) for the United States region 1, and 357 days (95% CrI: [334, 382]) for the United States region 9. The estimated standard deviations for the timing of peak transmission were 74 days (95% CrI: [45, 116]) for the Netherlands, 57 days (95% CrI: [48, 64]) for Denmark, 47 days (95% CrI: [31, 81]) in the United States region 1, and 33 (95% CrI: [20, 59]) in the United States region 9 (Table 1). Thus, the variation in peak timing in temperate regions (sd of about 35-60 days) is much smaller than the 100-day to 160-day variation in peak ILI+ seen in Vietnam.

As an alternate comparative approach, the posterior distribution of all peak timings can be constructed and compared between tropical and temperate areas, with peak season defined as the period encompassing 90% of the posterior density of φ, as described previously (Figure 5). In temperate zones, peak season is contained within a one-to-three-month time window whereas peak season in three regions of Vietnam would have to be defined as a 6-7 month period. Additionally, when separated out by subtype in southern and central Vietnam, the peak ILI+ timings do not overlap, indicating that there is no preferred climatic period or school-term period that favors higher levels of influenza transmission. In temperate zones, all seasons encompassed late fall and winter months and lasted between 56 and 116 days, with the seasons for the Netherlands, Denmark, US region 1, and US region 9 spanning 11 Nov – 3 Mar; 8 Dec – 3 Apr; 22 Sep – 15 Jan; and 14 Oct – 9 Dec, respectively (Table 1, Figure 5).

Other parameters were shown to differ among the model fits comparing Vietnam to temperate settings. The fits from the Netherlands and Denmark showed notably higher baseline transmission parameter values, but lower relative transmission increases during outbreak periods. The estimated duration of epidemics was also estimated to be notably higher for A/H1 and A/H3 in Vietnam compared to the other model fits. Sporadic introduction of cases occurred more frequently among ILI+, A/H1, and A/H3 in Vietnam compared to the other model fits. The estimates for duration of immunity after infection for ILI+ and influenza B in Vietnam were approximately half that from the other model fits (Table S2).

## 3. Discussion

This study aimed to address seasonality of influenza in Vietnam by determining its existence through a mathematical model able to test the hypothesis that seasonal repeatability does exist. Repeatability was defined here as the consistent and predictable generation of epidemics at the same time each year. Fitting the model to positive influenza incidence in temperate settings demonstrated the model’s ability to detect seasonality when it exists through parameters representing the timings of periods with increased transmission. Applying this model to positive influenza incidence in Vietnam, using combined incidence as well as for individual (sub)types, showed much weaker evidence of seasonality through more widely varying timings of increased transmission. While the four temperate locations examined in this study showed that peak transmission occurs consistently during winter months, with timings between peaks having low variance, all three regions of Vietnam show peak transmission occurring throughout the year, typically with high variance in the inferred timings of a true “peak” if it were to exist.

Assessments of seasonality are critical for tropical countries as they determine whether vaccine rollout can or should be timed to occur before an influenza season. The knowledge that regular cycles are weak or nonexistent is important for such preparedness efforts. In contrast to temperate locations, where vaccination and health messaging can effectively be targeted during autumn, there is not a clear time of year when vaccines can be prioritized in Vietnam for maximum protection. Because influenza can be seen year-round in Vietnam and other nearby tropical locations, identifying a single time of year to prioritize vaccination is less likely to be most effective, despite previous attempts based on data descriptions (39). Similarly, because the variance of timing between peaks is wide, knowledge of the timing of a current peak is not informative of the timing of the following peak. Currently, despite an existing vaccine policy aiming to target some at-risk demographics (49), influenza vaccination is not common in Vietnam even among some high-risk demographics, particularly in rural areas (50,51), and the results of this study highlight a major challenge in implementing a vaccination strategy.

### 3.1 Past consensus on influenza seasonality in the tropics

The results of this study showed very weak evidence of seasonal or nonannual cycles of influenza in Vietnam. This is consistent with some research investigating seasonality of tropical influenza. Previous studies have also noted a lack of seasonal trends in tropical locations, whether by acknowledging that patterns were not identified (52–54) or by describing seasons of influenza that do not show consistency in timing (37,55). Nevertheless, other studies have found evidence of regular annual (19,41,56,57) and nonannual (28,58) cycles in tropical influenza and have recommended preparedness efforts such as vaccination based on these. Similar to the results of this study, other studies identifying seasonal patterns have noted that the strength of seasonality in tropical locations is not as strong as that seen in temperate locations. Common seasonal patterns included yearly peaks occurring at different times of year (56) and observing two peaks per year (37,48,59).

While many previous works have noted that tropical locations can experience one or two epidemic peaks each year, the variability in their timings, as found in this study, show that they are not necessarily consistent or repeatable. By quantifying the comparatively large variance in timings of epidemic peaks in Vietnam (Figure 4), we show that knowledge of the timing of one epidemic peak does little to inform the timing of the following epidemic peak, and therefore is not repeatable.

Fitting the same model to data from different locations within Vietnam led to different posterior distributions for model parameters. This creates difficulty in inferring mechanisms of influenza due to the differences in inferred parameter values among different locations and different (sub)types. It is possible that other factors not accounted for in this model are important determinants of influenza dynamics in Vietnam. Under such circumstances, the estimated model parameters shown in Table S2 may represent these additional determinants of influenza burden. However, given the purposes of this study, this further shows that standard assumptions regarding influenza dynamics that are commonly applied to temperate locations are unlikely to directly apply in the tropics, indicating that mechanisms of influenza are likely to differ.

### 3.2. Differences in transmission intensity between temperate and tropical regions

A key comparison missing in the literature is whether the inherent community-level transmission of influenza virus (that is, one level beyond what is measured in household studies) is higher in tropical regions or temperate regions. Additionally, this question would need to be answered separately for peak transmission and baseline transmission periods. Measures of population density, crowding (60), or mobility (61) are likely the most appropriate proxies for community transmission levels that would be expected to be observed for any respiratory virus. Our inferred dynamics are consistent with higher crowding/aggregation/transmission levels in temperate areas, and also with temperate areas experiencing smaller changes in transmission amplitude. Our current study, however, is unable to identify this particular difference in inherent community-level transmission. Further work effectively comparing population mixing across the locations investigated in this study requires substantial resources and poses challenges in implementation.

The model showed close fits to the observed data from Vietnam as well as from the temperate locations, with the temperate locations showing closer and more precise fits (Figure 3). Across the different model fits among different (sub)types and locations, some parameters of the model showed different values. The fits from the four temperate countries showed transmission parameters that were higher than those estimated across (sub)types in Vietnam. The temperate transmission parameters, along with the assumed five-day duration of infection, are consistent with a reproduction number between two and three (Table S2). Previous estimates for the reproduction number in Europe and North America lie between one and two (62,63), though some estimates above two have been seen (64,65). The fits for Vietnam, however, suggested basic reproduction numbers below, but near, one, with effective reproduction numbers increasing to values above one during the fitted periods of increased infectivity. While reproduction numbers of influenza have previously been estimated to be above one in tropical settings and other countries in Asia, some evidence has shown that reproduction numbers globally trend higher in countries farther from the equator, and some tropical locations in Brazil have seen reproduction numbers below one (66–68). This may explain the seemingly constant presence of influenza in Vietnam; an effective reproduction number that is consistently close to one would allow infections to persist in the population in lower quantities.

Many of the model fits for Vietnam show trajectory shapes that are less symmetric than a typical epidemic curve (compare to epidemic curves see for the four temperate locations) particularly when fitting ILI+ (Fig 3). This behavior was examined and was found to result from the combination of low estimates for the baseline transmission parameter along with high amplitude of forcing. After epidemic periods begin, R_eff_ rises above one abruptly (this is an artefact of the model construction), allowing epidemic behavior to occur. After this period ends, R_eff_ falls below one, but there are both adequate proportions of the population infectious and susceptible to sustain a slow decline in infections rather than a quick decline. A higher baseline transmission parameter would lead to higher peaks in transmission, and therefore a steeper decline in transmission when epidemic periods end.

In contrast to the differences in fitted transmission parameters, the relative increases in transmission during epidemic periods were notably higher in Vietnam compared to temperate countries. While the United States, Denmark, and the Netherlands observed transmission increases of approximately ten percent, some model fits from Vietnam showed increases of over 50 percent. The amplitudes from both Vietnam and temperature locations, defined here as the product of this percentage increase and baseline transmission, differ somewhat from an estimated amplitude of 0.35 estimated during the 2009 H1N1 pandemic in the United States (69). Over such a short time, it is unlikely that there are true mechanistic reasons for an increase in transmission of nearly 50 percent; these large relative increases in transmission, as well as the timings of them denoted by the *φ*_*k*_ parameters, may instead represent irregular, less predictable events in Vietnam that can trigger an increase in transmission (e.g. the arrival of the virus to a more susceptible population pocket). This also relates to the lower baseline transmission rates in Vietnam; a high relative increase is more plausible when baseline transmission is low.

Even within the (sub)types across the different regions of Vietnam, differences in model parameters were observed. For example, small differences were seen in transmission and duration of immunity of subtype A/H1 across the three regions of Vietnam (Table S2). It is less likely that there are notable differences in virus characteristics across locations, but differences in transmission of the same (sub)type may result from different contact patterns or population densities. Similarly, epidemic-related parameters differed across locations such as timings of peak transmission, magnitude of transmission increase, and duration of transmission increase. These differences in epidemic characteristics are more likely to result from these differences in population densities and contacts across Vietnam. The observed differences are indicative of the wide heterogeneity in influenza dynamics in Vietnam and the lack of consistent seasonality; epidemic timing, amplitude, and duration are widely variable and unpredictable.

### 3.3. Importance of methodological approach

In this study, we used a mathematical model to measure repeatability to evaluate seasonality through peak timings, with wavelets and autocorrelation functions used for preliminary evidence. The use of wavelets has been common for addressing influenza seasonality in previous studies (19,41) but the use of mathematical models has not. Other works directly addressing seasonality in the tropics have used various types of statistical models, including linear and logistic regression models (37,52,58), general estimating equations (55), or clustering methods (53). Within these, it was common to incorporate sinusoidal functions to represent seasonal patterns (59), as we did with our time-varying transmission parameter. These non-mechanistic approaches provide evidence for or against annual patterns in time series, but they do not allow us to evaluate peak timings in a model that explicitly includes the mass-action dynamics, immune acquisition, and immune loss that is known to occur in influenza epidemics.

Rather than applying previously derived seasonal associations based on results observed in temperate regions to data from Vietnam, this study directly assessed whether seasonality exists in a human-to-human pathogen transmission system. This avoided the inherent assumption made in previous analyses that the associations found between seasonal factors such as climate and influenza risk are consistent across temperate and tropical locations. Our study also benefitted from the use of a model that estimated both peak timing and the variability of peak timings. This allowed us to use a unified inference system for temperate and tropical influenza, to validate that consistent winter peak timing was robustly inferred in all temperate data, and to compute the variability in timing of a hypothesized influenza season in the tropics.

This study relies on the assumption that the models applied to individual (sub)types of influenza in Vietnam function independently of each other. The model does not account for coinfection and cross-immunity among the three subtypes. This is a future extension of this work; a challenge in incorporating this was that it rarely allowed the subtypes to trade off while maintaining a single set of *φ*_*k*_ parameters. The aim of this study focused on identifying annual or nonannual cyclic trends, for which the *φ*_*k*_ parameters are critical.

Repeatable annual timing of influenza in the tropics, as evaluated by determining whether epidemics occur in regularly spaced time intervals in Vietnam, appears unlikely, as evidenced by an inferred high variance in peak time. Compared to temperate locations, three regions of Vietnam showed notably higher variation in when increased influenza transmission occurs. These findings suggest that there are no strong annual or nonannual cycles in influenza transmission, and timings of increased transmission are not primarily driven by cyclic or predictable phenomena. It follows that management and prevention of influenza in the tropics presents a challenging direction for public health research as future epidemics will be irregularly timed and difficult to predict.

## 4. Methods and materials

### 4.1 Influenza case data

In 2006, Vietnam’s National Institute of Hygiene and Epidemiology (NIHE) established a sentinel surveillance system for influenza, consisting of 16 hospitals throughout northern, central, and southern Vietnam, 15 of which provided sufficient data for this study. This syndromic surveillance system collected weekly counts of patients with influenza-like illness (ILI), the number of PCR-confirmed influenza cases, and the number of PCR tests administered each week (70,71). Among PCR-confirmed cases, the numbers of patients positive for subtype A/H1 (differentiating between 1977 lineage and pandemic 2009 lineage), subtype A/H3, and type B were included. Data were reported between 2006 and 2015, providing ten years of observation. Seven hospitals are located in northern Vietnam, including four in the city of Hanoi; four are located in central Vietnam; and four are located in southern Vietnam, including two in Ho Chi Minh City (Figure 1).

For the purposes of data analysis, we detrended ILI cases by dividing each weekly case count by the one-year weekly average of cases centered at that week. This can be viewed as a log-scale transformation of a z-score, with *ζ* = *e*^*z*^; this *ζ*-score approach preserves the relative sizes of case counts in neighboring weeks even after detrending, allowing for mechanistic fitting of a traditional epidemic incidence curve. This approach of detrending using a one-year moving average preserves any potential annual cycles present in the data while acting as a high-pass filter for trends greater than one year. We then multiplied the detrended ILI by the percentage of tests that were positive for each of the three subtypes to obtain a detrended positive ILI (ILI+) for each of three subtypes (72). Detrended ILI+ values for each hospital were then summed together within northern, central, and southern Vietnam, with each hospital weighted by the population of the province in which the hospital is located. One hospital in the southern Vietnamese city of Cai Be is located on the border of two provinces; therefore, its population weight was the average of the two province populations. Summing hospital totals minimizes the number of weeks with missing data. During weeks where some, but not all, hospitals provided data, the weighted average of ILI+ among hospitals presenting data were used. Separate analyses were run for subtype A/H1, subtype A/H3, type A, type B, and combined ILI+ for each of northern, central, and southern Vietnam.

We collected influenza surveillance data from the European Centers for Disease Control, which contain weekly case counts for all positive influenza between 2010 and 2019, but without specifying (sub)type. Data from the Netherlands and Denmark were the most complete data sets and therefore used in this study. We also collected weekly ILI cases for the United States (US) from the Centers for Disease Control and Prevention, which spanned between 2006 and 2015. We used data from two regions defined by the Department of Health and Human Services which are located on opposite sides of the US in order to increase variety of temperate locations. Region 1 spans the northeastern US, and region 9 encompasses the western US (73). We detrended data from the Netherlands, Denmark, and the US using the same *ζ*-score approach we used for the ILI+ data from Vietnam.

### 4.2. Patterns of seasonality

We conducted two standard time-series analyses to assess preliminary evidence of seasonality. The first consisted of an autocorrelation function (ACF), measuring lag periods for which the time series has high autocorrelation. Lag periods with high autocorrelation indicate a repeated pattern of that length throughout the data. The second consisted of a wavelet decomposition using a Morlet wavelet, which fits an oscillating curve with multiple inflection points, with varying periodicities, to the observed time series data. This analysis reveals not only cycle lengths, but where within the time series these cycles can be found. The results of this analysis show where along the time series this function can fit the data given different period lengths. A particular periodicity fitting consistently over the entire time span provides evidence of stationarity, which can be interpreted as evidence of cyclic patterns.

### 4.3. Measuring repeatability

We developed an ordinary differential equations model to identify and quantify repeatability of influenza. Using a mathematical model to measure repeatability allows us (*i*) to include the natural mass-action dynamics of respiratory virus transmission and (*ii*) to add peak periods into these natural dynamics that can later be characterized as repeatable or not. The model takes the form of a SIRS model, which includes a sinusoidal time-varying transmission parameter, *β*(*t*), that incorporates timings of potential seasons (74,75). We define the model by

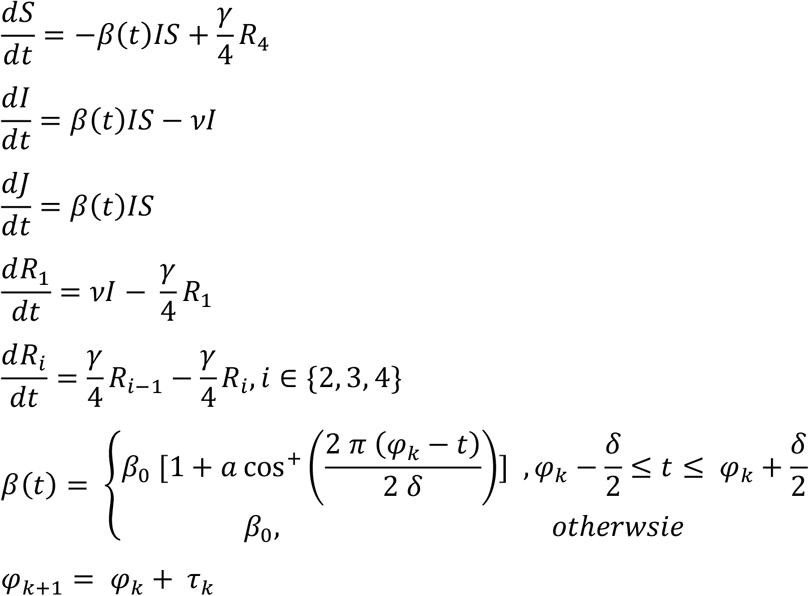

where cos^+^ is the positive-valued cosine function, *φ*_*k*_ represent timings of peak transmission, and *τ*_*k*_ represent the time (in days) between *φ*_*k*_ and *φ*_*k*+1_. The *τ*_*k*_ parameters are free and independent of each other to allow for completely irregular and non-annual epidemics. It follows that, while the *φ*_*k*_ values depend on each other in that *φ*_*k*+1_ > *φ*_*k*_ by definition, the distance between the two values is independent of either value. The number of *φ*_*k*_ ‘peak timing’ parameters (typically between 8 and 14 in our 10-year data set) was chosen a priori for each location and subtype based on visual examination of incidence trends followed by comparing preliminary model fits with different numbers of *φ*_*k*_ parameters. The three models with 13 or 14 *φ*_*k*_ were those for all ILI+ in northern, central, and southern Vietnam. Half (6/12) of the models for (sub)types in Vietnam and all four temperate models used 10 *φ*_*k*_ parameters. In the model, *δ* represents the duration, in days, of increased transmission (i.e. the duration of the increased transmission), *γ* represents the mean rate of waning immunity (reciprocal of duration of immunity in days), and *a* represents the relative increase in transmission, where *aβ*_0_ is the amplitude of the cosine function. The locations of the *φ*_*k*_ parameters (if consistent and repeatable) would correspond to annual seasonal dynamics. Regularly spaced peaks (i.e. low variance of timings between peaks) indicate strong annual seasonality, and higher variance of timings between peaks indicates weak annual seasonality.

The model also incorporates multiple recovery classes in order to reduce the variance in recovery time (76). Four recovery classes were chosen. In our data, when examining individual subtype data, periods of zero ILI+ exist. In order to facilitate time periods where zero prevalence occurs, a model behavior was added outside of the differential equations where ILI+ is set to zero based on a parameter *z* (fit from the data) at which point all infected individuals immediately recover. When ILI+ falls below *z*, ILI+ is set to zero. To then avoid extinction in the model, an immigration parameter was added that spontaneously inserts an influenza case from the susceptible population every *n* days (fit from the data). This represents influenza transmission occurring outside of the study area and then being introduced into the study area.

### 4.4. Model fitting

To fit the model to observed data, we ran the model for a ten-year period as burn-in followed by a second ten-year period for model fitting and comparison to observed data. Cumulative incidence variables (*J*) defined in the model were used to construct the model’s weekly incidence Δ*J*. We multiplied modeled ILI+ by an estimated reporting parameter, *ρ*, and then fit the output to observed data through a normal likelihood with fixed variance. Selection of a normal likelihood with fixed variance is a sum of squared errors fit, directly comparing the observed and predicted values. We also estimated the variance of the *τ*_*k*_ by estimating parameters representing their mean *μ*_*τ*_ and standard deviation *σ*_*τ*_ from a normal distribution and comparing all *τ*_*k*_ to this normal distribution in the likelihood fitting. This standard deviation is an important parameter for assessing strength of seasonality; low standard deviation of *τ*_*k*_ (and thus of *φ*_*k*_) provides evidence of strong seasonality because it indicates that the *φ*_*k*_ repeat at the same time every year.

We fit the model to data for each of subtypes A/H1 and A/H3, combined type A, type B, and all ILI+ in northern, central, and southern Vietnam using Markov Chain Monte Carlo (MCMC) sampling with the Metropolis-Hastings algorithm to estimate parameters. We also fit the model to ILI+ data from the Netherlands and Denmark as well as ILI data from the US to demonstrate the ability of the model to detect seasonal patterns in temperate countries. Parameters estimated include the transmission parameter (β_0_), mean duration of immunity (*γ*^−1^), magnitude of increased transmission (*a*), duration of increased transmission (δ), timings of epidemic periods (*φ*_*k*_), incidence level when epidemics end (*z*), and frequency of case importation (*n*). We selected diffuse priors (Table S1). Assumed parameters include duration of illness (five days) (77).

The model we developed is sensitive to initial parameter values and can produce different outcomes through small changes. For example, holding other parameters constant and modifying the transmission parameter by 10% can lead to very different trajectories in incidence. Therefore, we conducted wide parameter space searches followed by close examination of changes in individual parameters to inform starting values prior to running the MCMC chains. This allowed smaller variances to be selected for the proposal distributions to increase the probability of a proposal being accepted (Table S1). To balance exploring parameter space during MCMC and efficient sampling, variance from proposal distribution was modified after every 50,000 attempted iterations to shrink it if there were too few accepted samples or expand if there were too many accepted samples. If between 10 and 90 percent of the last 50,000 attempted iterations were accepted, their variance was used to update the proposal distribution (78). Otherwise, the variance of the proposal distribution was increased or decreased by 10 percent. Doing so allowed the acceptance rate of the MCMC to reach approximately five percent after seeing a starting acceptance rate below one percent.

We ran the MCMC for four independent chains for each of the five models using data from northern, central, and southern Vietnam as well as the models using data from the Netherlands, Denmark, and the US. A minimum of 50,000 burn-in samples were run, and then convergence was evaluated using visual inspection of trace plots and the Gelman-Rubin statistic. After convergence was reached, an additional 50,000 samples were collected to generate posterior distributions, thinning to include every fifth sample. Results are presented from the chain with the highest median posterior likelihood value, with results from other chains used as robustness validation. Median values for each parameter with 95% credible intervals (CrI) are presented. The MCMC inference was run using R version 4.0.3 (79), using the ‘mvtnorm’ package for drawing parameters from the proposal distribution (80).

## Data Availability

All relevant data and code are available at github.com/jlservadio/Influenza_Seas_VN

## Data Availability

All relevant data and code are available at github.com/jlservadio/Influenza_Seas_VN

## Supplement

**Figure S1**. Autocorrelation functions for ten years of weekly influenza incidence. Columns from left to right show autocorrelation plots for northern Vietnam, central Vietnam, southern Vietnam, and four temperate locations.

**Figure S2**. Morlet wavelet decompositions for ten years of weekly influenza incidence data. Columns from left to right show results for northern Vietnam, central Vietnam, southern Vietnam, and temperate regions.

**Table S1**. Prior distributions and standard deviations for proposal distributions for all parameters estimated through MCMC. The row for tau_k indicates that all tau parameters were given the same prior distribution and standard deviation for the proposal distribution.

**Table S2**. Posterior estimates with 95% credible intervals for all estimated parameters from all models.

## Acknowledgements

The authors thank the National Institute of Hygiene and Epidemiology in Vietnam for providing the data used in this study. The authors also thank the leadership and staff at the sentinel hospitals that collected the data used as well as Ephraim Hanks and Ottar Bjørnstad for providing methodological and substantive feedback on this work.

The authors acknowledge the resources of Pennsylvania State University’s Institute for Computational and Data Sciences Roar supercomputer for data analysis and management.

## Author contributions

Conceptualization: JLS, MFB

Investigation: PQT, MFB

Methodology: JLS, MC, MFB

Software: JLS, MFB

Formal Analysis: JLS

Visualization: JLS, MC, MFB

Supervision: MFB

Funding acquisition: JLS, MFB

Writing – Original draft: JLS

Writing – Review and Editing: JLS, PQT, MC, MFB

## Data Availability

All relevant data and code are available at github.com/jlservadio/Influenza_Seas_VN

## References

1. Peteranderl C, Herold S, Schmoldt C. Human Influenza Virus Infections. Semin Respir Crit Care Med. 2016 Aug;37(4):487–500.

2. World Health Organization. Influenza [Internet]. 2022 [cited 2022 Apr 19]. Available from: https://www.who.int/teams/health-product-policy-and-standards/standards-and-specifications/vaccines-quality/influenza

3. Paget J, Spreeuwenberg P, Charu V, Taylor RJ, Iuliano AD, Bresee J, et al. Global mortality associated with seasonal influenza epidemics: New burden estimates and predictors from the GLaMOR Project. J Glob Health [Internet]. [cited 2021 Feb 1];9(2). Available from: https://www.ncbi.nlm.nih.gov/pmc/articles/PMC6815659/

4. Iuliano AD, Roguski KM, Chang HH, Muscatello DJ, Palekar R, Tempia S, et al. Estimates of global seasonal influenza-associated respiratory mortality: a modelling study. The Lancet. 2018 Mar 31;391(10127):1285–300.

5. World Health Organization. Up to 650 000 people die of respiratory diseases linked to seasonal flu each year [Internet]. 2017 [cited 2021 Feb 6]. Available from: https://www.who.int/news/item/13-12-2017-up-to-650-000-people-die-of-respiratory-diseases-linked-to-seasonal-flu-each-year

6. Taubenberger JK, Morens DM. The Pathology of Influenza Virus Infections. Annu Rev Pathol. 2008;3:499–522.

7. Wu P, Presanis AM, Bond HS, Lau EHY, Fang VJ, Cowling BJ. A joint analysis of influenza-associated hospitalizations and mortality in Hong Kong, 1998–2013. Sci Rep. 2017 Apr 20;7(1):929.

8. Molinari NAM, Ortega-Sanchez IR, Messonnier ML, Thompson WW, Wortley PM, Weintraub E, et al. The annual impact of seasonal influenza in the US: measuring disease burden and costs. Vaccine. 2007 Jun 28;25(27):5086–96.

9. Monto AS, Fukuda K. Lessons From Influenza Pandemics of the Last 100 Years. Clin Infect Dis Off Publ Infect Dis Soc Am. 2020 Mar 1;70(5):951–7.

10. Paget J, Marquet R, Meijer A, van der Velden K. Influenza activity in Europe during eight seasons (1999–2007): an evaluation of the indicators used to measure activity and an assessment of the timing, length and course of peak activity (spread) across Europe. BMC Infect Dis. 2007 Nov 30;7(1):141.

11. Lofgren E, Fefferman NH, Naumov YN, Gorski J, Naumova EN. Influenza seasonality: underlying causes and modeling theories. J Virol. 2007 Jun;81(11):5429–36.

12. Byambasuren S, Paradowska-Stankiewicz I, Brydak LB. Epidemic Influenza Seasons from 2008 to 2018 in Poland: A Focused Review of Virological Characteristics. Trends Biomed Res. 2020 Jan 28;1251:115–21.

13. Redondo-Bravo L, Delgado-Sanz C, Oliva J, Vega T, Lozano J, Larrauri A. Transmissibility of influenza during the 21st-century epidemics, Spain, influenza seasons 2001/02 to 2017/18. Eurosurveillance. 2020 May 28;25(21):1900364.

14. McAndrew T, Reich NG. Adaptively stacking ensembles for influenza forecasting. Stat Med. 2021 Dec 30;40(30):6931–52.

15. Midgley CM, Haynes AK, Baumgardner JL, Chommanard C, Demas SW, Prill MM, et al. Determining the Seasonality of Respiratory Syncytial Virus in the United States: The Impact of Increased Molecular Testing. J Infect Dis. 2017 Aug 1;216(3):345–55.

16. Killerby ME, Biggs HM, Haynes A, Dahl RM, Mustaquim D, Gerber SI, et al. Human coronavirus circulation in the United States 2014–2017. J Clin Virol. 2018 Apr 1;101:52–6.

17. Shah MM, Winn A, Dahl RM, Kniss KL, Silk BJ, Killerby ME. Seasonality of Common Human Coronaviruses, United States, 2014–20211. Emerg Infect Dis. 2022 Oct;28(10):1970–6.

18. Morikawa S, Kohdera U, Hosaka T, Ishii K, Akagawa S, Hiroi S, et al. Seasonal variations of respiratory viruses and etiology of human rhinovirus infection in children. J Clin Virol Off Publ Pan Am Soc Clin Virol. 2015 Dec;73:14–9.

19. Thai PQ, Choisy M, Duong TN, Thiem VD, Yen NT, Hien NT, et al. Seasonality of absolute humidity explains seasonality of influenza-like illness in Vietnam. Epidemics. 2015 Dec 1;13:65–73.

20. Luca GD, Kerckhove KV, Coletti P, Poletto C, Bossuyt N, Hens N, et al. The impact of regular school closure on seasonal influenza epidemics: a data-driven spatial transmission model for Belgium. BMC Infect Dis. 2018 Jan 10;18:29.

21. Simpson RB, Alarcon Falconi TM, Venkat A, Chui KHH, Navidad J, Naumov YN, et al. Incorporating calendar effects to predict influenza seasonality in Milwaukee, Wisconsin. Epidemiol Infect. 2019 Sep 11;147:e268.

22. Lowen AC, Mubareka S, Steel J, Palese P. Influenza Virus Transmission Is Dependent on Relative Humidity and Temperature. PLOS Pathog. 2007 Oct 19;3(10):e151.

23. Shaman J, Kohn M. Absolute humidity modulates influenza survival, transmission, and seasonality. Proc Natl Acad Sci. 2009 Mar 3;106(9):3243–8.

24. Tamerius J, Nelson MI, Zhou SZ, Viboud C, Miller MA, Alonso WJ. Global influenza seasonality: reconciling patterns across temperate and tropical regions. Environ Health Perspect. 2011 Apr;119(4):439–45.

25. Moriyama M, Hugentobler WJ, Iwasaki A. Seasonality of Respiratory Viral Infections. Annu Rev Virol. 2020 Sep 29;7(1):83–101.

26. Fares A. Factors Influencing the Seasonal Patterns of Infectious Diseases. Int J Prev Med. 2013 Feb;4(2):128–32.

27. Feng S, Chiu SS, Chan ELY, Kwan MYW, Wong JSC, Leung CW, et al. Effectiveness of influenza vaccination on influenza-associated hospitalisations over time among children in Hong Kong: a test-negative case-control study. Lancet Respir Med. 2018 Dec;6(12):925–34.

28. Tamerius JD, Shaman J, Alonso WJ, Bloom-Feshbach K, Uejio CK, Comrie A, et al. Environmental Predictors of Seasonal Influenza Epidemics across Temperate and Tropical Climates. PLOS Pathog. 2013 Mar 7;9(3):e1003194.

29. Viboud C, Alonso WJ, Simonsen L. Influenza in Tropical Regions. PLoS Med [Internet]. 2006 Apr [cited 2021 Feb 12];3(4). Available from: https://www.ncbi.nlm.nih.gov/pmc/articles/PMC1391975/

30. Gentile A, Paget J, Bellei N, Torres JP, Vazquez C, Laguna-Torres VA, et al. Influenza in Latin America: A report from the Global Influenza Initiative (GII). Vaccine. 2019 May 6;37(20):2670–8.

31. Gessner BD, Shindo N, Briand S. Seasonal influenza epidemiology in sub-Saharan Africa: a systematic review. Lancet Infect Dis. 2011 Mar 1;11(3):223–35.

32. Chadha MS, Potdar VA, Saha S, Koul PA, Broor S, Dar L, et al. Dynamics of influenza seasonality at sub-regional levels in India and implications for vaccination timing. PloS One. 2015;10(5):e0124122.

33. Cowling BJ, Caini S, Chotpitayasunondh T, Djauzi S, Gatchalian SR, Huang QS, et al. Influenza in the Asia-Pacific region: Findings and recommendations from the Global Influenza Initiative. Vaccine. 2017 Feb 7;35(6):856–64.

34. Muscatello DJ. Redefining influenza seasonality at a global scale and aligning it to the influenza vaccine manufacturing cycle: A descriptive time series analysis. J Infect. 2019 Feb 1;78(2):140–9.

35. Suntronwong N, Vichaiwattana P, Klinfueng S, Korkong S, Thongmee T, Vongpunsawad S, et al. Climate factors influence seasonal influenza activity in Bangkok, Thailand. PloS One. 2020;15(9):e0239729.

36. Liu Z, Zhang J, Zhang Y, Lao J, Liu Y, Wang H, et al. Effects and interaction of meteorological factors on influenza: Based on the surveillance data in Shaoyang, China. Environ Res. 2019 May;172:326–32.

37. Hirve S, Newman LP, Paget J, Azziz-Baumgartner E, Fitzner J, Bhat N, et al. Influenza Seasonality in the Tropics and Subtropics – When to Vaccinate? PLoS ONE [Internet]. 2016 Apr 27 [cited 2021 Feb 25];11(4). Available from: https://www.ncbi.nlm.nih.gov/pmc/articles/PMC4847850/

38. Simmerman JM, Chittaganpitch M, Levy J, Chantra S, Maloney S, Uyeki T, et al. Incidence, seasonality and mortality associated with influenza pneumonia in Thailand: 2005-2008. PloS One. 2009 Nov 11;4(11):e7776.

39. Saha S, Chadha M, Al Mamun A, Rahman M, Sturm-Ramirez K, Chittaganpitch M, et al. Influenza seasonality and vaccination timing in tropical and subtropical areas of southern and south-eastern Asia. Bull World Health Organ. 2014 May 1;92(5):318–30.

40. Kosasih H, Roselinda null, Nurhayati null, Klimov A, Xiyan X, Lindstrom S, et al. Surveillance of influenza in Indonesia, 2003–2007. Influenza Other Respir Viruses. 2013 May;7(3):312–20.

41. Alonso WJ, Viboud C, Simonsen L, Hirano EW, Daufenbach LZ, Miller MA. Seasonality of influenza in Brazil: a traveling wave from the Amazon to the subtropics. Am J Epidemiol. 2007 Jun 15;165(12):1434–42.

42. Deyle ER, Maher MC, Hernandez RD, Basu S, Sugihara G. Global environmental drivers of influenza. Proc Natl Acad Sci U S A. 2016 Nov 15;113(46):13081–6.

43. Kamigaki T, Chaw L, Tan AG, Tamaki R, Alday PP, Javier JB, et al. Seasonality of Influenza and Respiratory Syncytial Viruses and the Effect of Climate Factors in Subtropical-Tropical Asia Using Influenza-Like Illness Surveillance Data, 2010-2012. PloS One. 2016;11(12):e0167712.

44. Zhang Y, Ye C, Yu J, Zhu W, Wang Y, Li Z, et al. The complex associations of climate variability with seasonal influenza A and B virus transmission in subtropical Shanghai, China. Sci Total Environ. 2020 Jan 20;701:134607.

45. Chong KC, Lee TC, Bialasiewicz S, Chen J, Smith DW, Choy WSC, et al. Association between meteorological variations and activities of influenza A and B across different climate zones: a multi-region modelling analysis across the globe. J Infect. 2020 Jan;80(1):84–98.

46. Ma P, Tang X, Zhang L, Wang X, Wang W, Zhang X, et al. Influenza A and B outbreaks differed in their associations with climate conditions in Shenzhen, China. Int J Biometeorol. 2022 Jan;66(1):163–73.

47. Nakapan S, Tripathi NK, Tipdecho T, Souris M. Spatial diffusion of influenza outbreak-related climate factors in Chiang Mai Province, Thailand. Int J Environ Res Public Health. 2012 Oct 24;9(11):3824–42.

48. Yuan H, Kramer SC, Lau EHY, Cowling BJ, Yang W. Modeling influenza seasonality in the tropics and subtropics. PLoS Comput Biol. 2021 Jun;17(6):e1009050.

49. Morales KF, Brown DW, Dumolard L, Steulet C, Vilajeliu A, Ropero Alvarez AM, et al. Seasonal influenza vaccination policies in the 194 WHO Member States: The evolution of global influenza pandemic preparedness and the challenge of sustaining equitable vaccine access. Vaccine X. 2021 Apr 20;8:100097.

50. Le XTT, Nguyen HT, Le HT, Do TTT, Nguyen TH, Vu LG, et al. Rural-urban differences in preferences for influenza vaccination among women of childbearing age: implications for local vaccination service implementation in Vietnam. Trop Med Int Health TM IH. 2021 Feb;26(2):228–36.

51. Nguyen TTM, Lafond KE, Nguyen TX, Tran PD, Nguyen HM, Ha VTC, et al. Acceptability of seasonal influenza vaccines among health care workers in Vietnam in 2017. Vaccine. 2020 Feb 18;38(8):2045–50.

52. Bloom-Feshbach K, Alonso WJ, Charu V, Tamerius J, Simonsen L, Miller MA, et al. Latitudinal variations in seasonal activity of influenza and respiratory syncytial virus (RSV): a global comparative review. PloS One. 2013;8(2):e54445.

53. Alonso WJ, Guillebaud J, Viboud C, Razanajatovo NH, Orelle A, Zhou SZ, et al. Influenza seasonality in Madagascar: the mysterious African free-runner. Influenza Other Respir Viruses. 2015 May;9(3):101–9.

54. Li Y, Reeves RM, Wang X, Bassat Q, Brooks WA, Cohen C, et al. Global patterns in monthly activity of influenza virus, respiratory syncytial virus, parainfluenza virus, and metapneumovirus: a systematic analysis. Lancet Glob Health. 2019 Aug;7(8):e1031–45.

55. Gordon A, Ortega O, Kuan G, Reingold A, Saborio S, Balmaseda A, et al. Prevalence and Seasonality of Influenza-like Illness in Children, Nicaragua, 2005–2007. Emerg Infect Dis. 2009 Mar;15(3):408–14.

56. Koul PA, Broor S, Saha S, Barnes J, Smith C, Shaw M, et al. Differences in influenza seasonality by latitude, northern India. Emerg Infect Dis. 2014 Oct;20(10):1723–6.

57. Moura FEA, Perdigão ACB, Siqueira MM. Seasonality of Influenza in the Tropics: A Distinct Pattern in Northeastern Brazil. Am J Trop Med Hyg. 2009 Jul 1;81(1):180–3.

58. Lam HM, Wesolowski A, Hung NT, Nguyen TD, Nhat NTD, Todd S, et al. Nonannual seasonality of influenza-like illness in a tropical urban setting. Influenza Other Respir Viruses. 2018;12(6):742–54.

59. Yu H, Alonso WJ, Feng L, Tan Y, Shu Y, Yang W, et al. Characterization of Regional Influenza Seasonality Patterns in China and Implications for Vaccination Strategies: Spatio-Temporal Modeling of Surveillance Data. PLoS Med. 2013 Nov 19;10(11):e1001552.

60. Dalziel BD, Kissler S, Gog JR, Viboud C, Bjørnstad ON, Metcalf CJE, et al. Urbanization and humidity shape the intensity of influenza epidemics in U.S. cities. Science. 2018;362(6410):75–9.

61. Charu V, Zeger S, Gog J, Bjørnstad ON, Kissler S, Simonsen L, et al. Human mobility and the spatial transmission of influenza in the United States. PLoS Comput Biol. 2017 Feb 10;13(2):e1005382.

62. Bettencourt LMA, Ribeiro RM. Real Time Bayesian Estimation of the Epidemic Potential of Emerging Infectious Diseases. PLoS ONE. 2008 May 14;3(5):e2185.

63. Smieszek T, Balmer M, Hattendorf J, Axhausen KW, Zinsstag J, Scholz RW. Reconstructing the 2003/2004 H3N2 influenza epidemic in Switzerland with a spatially explicit, individual-based model. BMC Infect Dis. 2011 May 9;11:115.

64. Truscott J, Fraser C, Cauchemez S, Meeyai A, Hinsley W, Donnelly CA, et al. Essential epidemiological mechanisms underpinning the transmission dynamics of seasonal influenza. J R Soc Interface. 2012 Feb 7;9(67):304–12.

65. Nikbakht R, Baneshi MR, Bahrampour A. Estimation of the Basic Reproduction Number and Vaccination Coverage of Influenza in the United States (2017-18). J Res Health Sci. 2018 Sep 22;18(4):e00427.

66. Chowell G, Viboud C, Simonsen L, Miller M, Alonso WJ. The reproduction number of seasonal influenza epidemics in Brazil, 1996–2006. Proc R Soc B Biol Sci. 2010 Jun 22;277(1689):1857–66.

67. Yamauchi T, Takeuchi S, Yamano Y, Kuroda Y, Nakadate T. Estimation of the effective reproduction number of influenza based on weekly reports in Miyazaki Prefecture. Sci Rep. 2019 Feb 22;9:2539.

68. Thompson R, Wood JG, Tempia S, Muscatello DJ. Global variation in early epidemic growth rates and reproduction number of seasonal influenza. Int J Infect Dis IJID Off Publ Int Soc Infect Dis. 2022 Sep;122:382–8.

69. Towers S, Feng Z. Pandemic H1N1 influenza: predicting the course of a pandemic and assessing the efficacy of the planned vaccination programme in the United States. Euro Surveill Bull Eur Sur Mal Transm Eur Commun Dis Bull. 2009 Oct 15;14(41):19358.

70. Nguyen HT, Dharan NJ, L. MTQ, Nguyen NB, Nguyen CT, Hoang DV, et al. National influenza surveillance in Vietnam, 2006-2007. Vaccine. 2009 Dec 11;28(2):398–402.

71. Nguyen YT, Graitcer SB, Nguyen TH, Tran DN, Pham TD, L. MTQ, et al. National surveillance for influenza and influenza-like illness in Vietnam, 2006–2010. Vaccine. 2013 Sep 13;31(40):4368–74.

72. Shaman J, Karspeck A, Yang W, Tamerius J, Lipsitch M. Real-Time Influenza Forecasts during the 2012–2013 Season. Nat Commun. 2013;4:2837.

73. Department of Health and Human Services. HHS Regional Offices [Internet]. HHS.gov. 2006 [cited 2022 Sep 22]. Available from: https://www.hhs.gov/about/agencies/iea/regional-offices/index.html

74. Hickmann KS, Fairchild G, Priedhorsky R, Generous N, Hyman JM, Deshpande A, et al. Forecasting the 2013-2014 influenza season using Wikipedia. PLoS Comput Biol. 2015 May;11(5):e1004239.

75. Papst I, Earn DJD. Invariant predictions of epidemic patterns from radically different forms of seasonal forcing. J R Soc Interface. 2019 Jul 26;16(156):20190202.

76. Wikle NB, Tran TNA, Gentilesco B, Leighow SM, Albert E, Strong ER, et al. SARS-CoV-2 epidemic after social and economic reopening in three U.S. states reveals shifts in age structure and clinical characteristics. Sci Adv. 2022 Jan 28;8(4):eabf9868.

77. Hien TT, Boni MF, Bryant JE, Ngan TT, Wolbers M, Nguyen TD, et al. Early pandemic influenza (2009 H1N1) in Ho Chi Minh City, Vietnam: a clinical virological and epidemiological analysis. PLoS Med. 2010 May 18;7(5):e1000277.

78. Shaby BA, Wells MT. Exploring an adaptive Metropolis algorithm. Tech Rep. 2010;1011–4.

79. R Core Team. R: A language and environment for statistical computing [Internet]. Vienna, Austria: R Foundation for Statistical Computing; 2020. Available from: https://www.R-project.org

80. Genz A, Bretz F, Miwa T, Mi X, Leish F, Sheipl F, et al. mvtnorm: Multivariate Normal and t Distributions [Internet]. 2021. Available from: https://CRAN.R-project.org/package=mvtnorm

