## Supplement for "Repeatability and timing of tropical influenza epidemics"

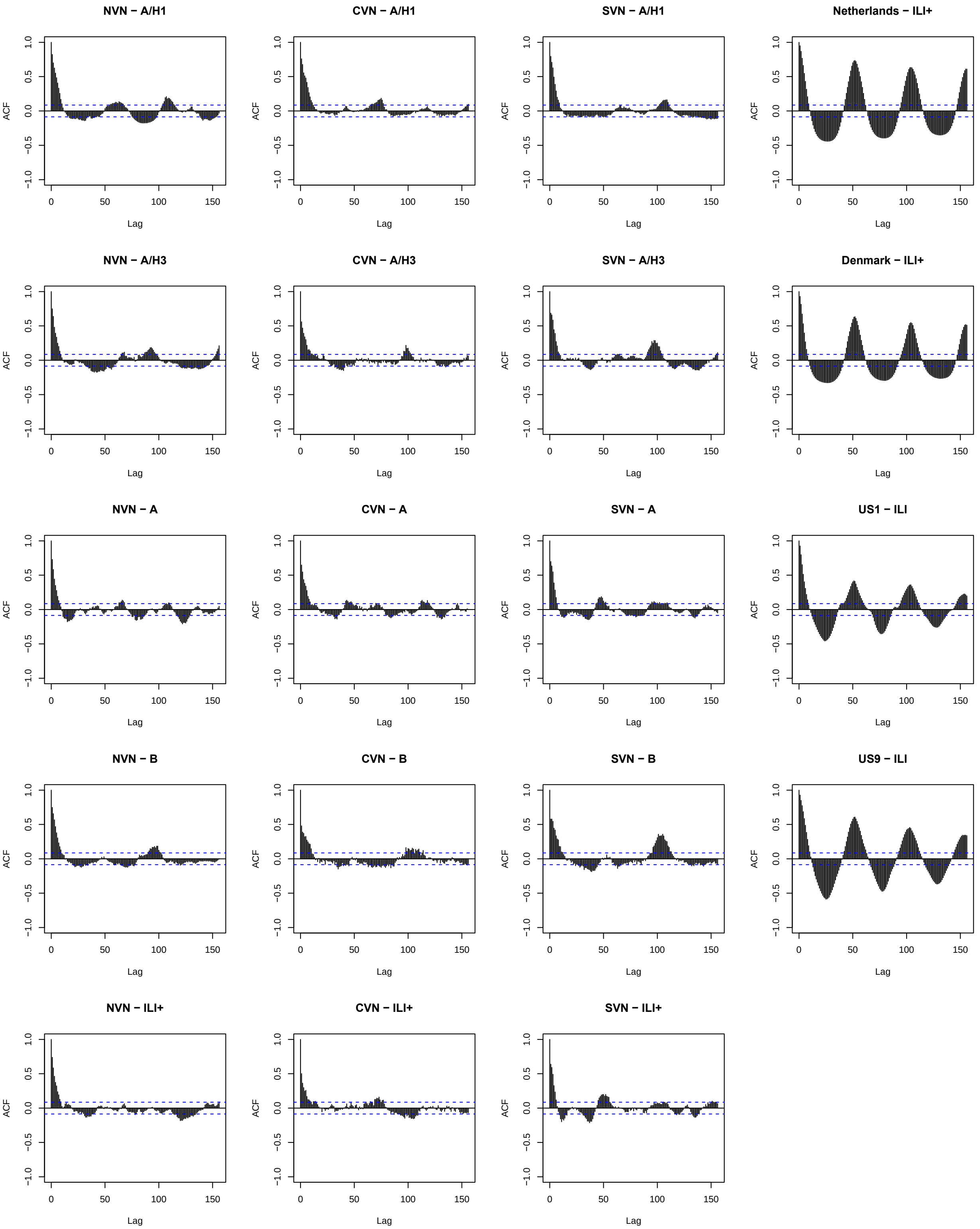


**Figure S1.** Autocorrelation functions for ten years of weekly influenza incidence. Columns from left to right show autocorrelation plots for northern Vietnam, central Vietnam, southern Vietnam, and four temperate locations.

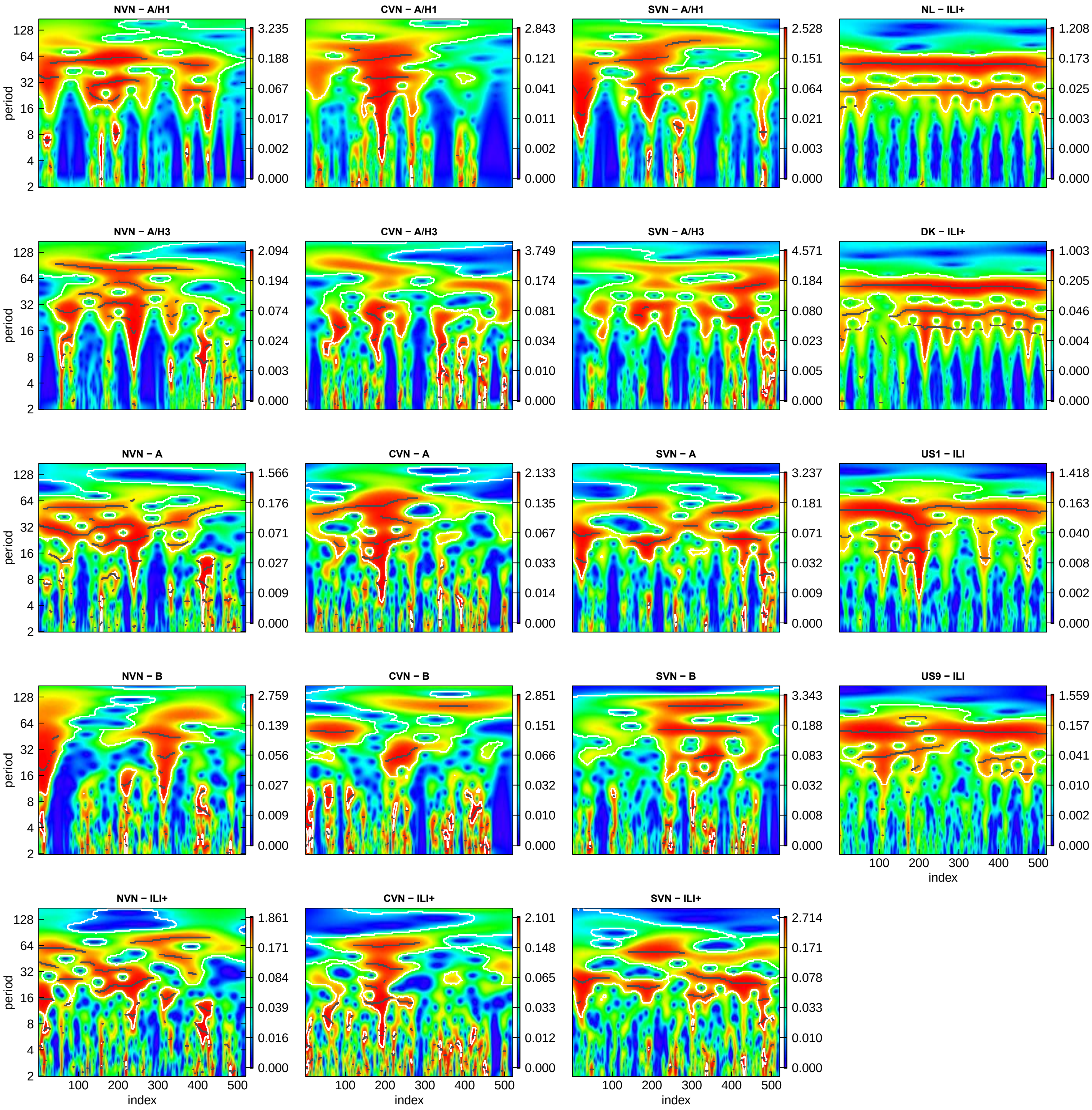


**Figure S2.** Morlet wavelet decompositions for ten years of weekly influenza incidence data. Columns from left to right show results for northern Vietnam, central Vietnam, southern Vietnam, and temperate regions.

**Table S1.** Prior distributions and standard deviations for proposal distributions for all parameters estimated through MCMC. The row for tau_k indicates that all tau parameters were given the same prior distribution and standard deviation for the proposal distribution.

| Parameter | Prior distribution | Proposal distribution standard deviation |
|---|---|---|
| beta | U(0.14, 1) | 0.01 |
| amp | Exp(1) | 0.01 |
| 1/gamma | U(180, 365x6) | 10 |
| phi.1 | U(0, 450) | 5 |
| tau_k | U(20, 700) | 5 |
| mu.tau | U(20, 700) | 5 |
| sigma.tau | U(1, 200) | 2 |
| rho (reporting) | U(0.0001, 0.1) | 0.0002 |
| delta (epidemic duration) | U(20, 180) | 2 |
| z (epidemic end) | U(0, 1000) | 5 |
| n (importation) | U(30, 1000) | 5 |

**Table S2.** Posterior estimates with 95% credible intervals for all estimated parameters from all models.

| param | Estimate_nvn_h1 | CrI_nvn_h1 | Estimate_nvn_h3 | CrI_nvn_h3 | Estimate_nvn_a | CrI_nvn_a |
|---|---|---|---|---|---|---|
| beta | 0.191 | [0.182, 0.199] | 0.193 | [0.186, 0.2] | 0.185 | [0.182, 0.187] |
| a | 0.865 | [0.762, 0.972] | 1.1 | [1.017, 1.164] | 0.319 | [0.269, 0.38] |
| gamma | 1167.179 | [622.615, 2140.566] | 1235.921 | [919.138, 1980.341] | 1198.055 | [429.296, 1948.444] |
| phi.1 | 117.76 | [112.107, 123.515] | 350.318 | [344.595, 355.679] | 132.857 | [126.557, 141.005] |
| tau.1 | 228.039 | [193.738, 249.014] | 183.662 | [175.847, 190.424] | 408.227 | [399.385, 416.61] |
| tau.2 | 544.771 | [521.626, 579.143] | 306.305 | [287.323, 324.288] | 365.815 | [359.76, 370.926] |
| tau.3 | 424.368 | [415.055, 433.565] | 323.493 | [304.244, 343.704] | 258.288 | [247.398, 272.063] |
| tau.4 | 482.182 | [467.075, 496.312] | 471.306 | [463.705, 477.86] | 160.353 | [147.496, 171.377] |
| tau.5 | 227.843 | [194.706, 249.651] | 655.038 | [650.428, 659.727] | 317.289 | [308.34, 325.374] |
| tau.6 | 530.439 | [510.884, 561.117] | 562.076 | [533.357, 569.091] | 173.031 | [162.386, 183.116] |
| tau.7 | 364.014 | [356.065, 371.441] | 155.903 | [148.26, 185.453] | 478.21 | [468.331, 488.918] |
| tau.8 | 412.559 | [398.475, 423.991] | 370.596 | [366.437, 373.829] | 275.044 | [265.083, 284.228] |
| tau.9 | NA | NA | NA | NA | 303.618 | [294.799, 313.588] |
| tau.10 | NA | NA | NA | NA | NA | NA |
| tau.11 | NA | NA | NA | NA | NA | NA |
| tau.12 | NA | NA | NA | NA | NA | NA |
| tau.13 | NA | NA | NA | NA | NA | NA |
| mu.tau | 398.891 | [306.572, 495.315] | 379.681 | [268.893, 498.257] | 307.318 | [224.729, 390.114] |
| sigma.tau | 133.839 | [82.882, 190.971] | 163.72 | [117.08, 196.448] | 116.795 | [71.974, 180.162] |
| rho | 0 | [0, 0] | 0 | [0, 0] | 0.063 | [0.049, 0.078] |
| delta | 88.82 | [77.9, 98.602] | 81.174 | [75.986, 86.706] | 73.468 | [62.262, 86.313] |
| z | 248.446 | [87.976, 537.422] | 517.496 | [333.328, 680.347] | 2.424 | [1.718, 3.246] |
| n | 31.508 | [31.028, 31.963] | 42.483 | [42.031, 42.956] | 31.479 | [31.038, 31.968] |

Table S2 cont.

| param | Estimate_nvn_b | CrI_nvn_b | Estimate_nvn_ili+ | CrI_nvn_ili+ |
|---|---|---|---|---|
| beta | 0.198 | [0.196, 0.2] | 0.189 | [0.188, 0.19] |
| a | 0.538 | [0.332, 0.983] | 0.332 | [0.243, 0.408] |
| gamma | 408.138 | [369.359, 488.017] | 1161.694 | [434.614, 1712.991] |
| phi.1 | 37.895 | [31.405, 44.396] | 50.225 | [47.141, 54.7] |
| tau.1 | 399.457 | [334.645, 504.444] | 133.653 | [126.755, 139.791] |
| tau.2 | 238.835 | [126.457, 307.975] | 366.905 | [357.764, 374.929] |
| tau.3 | 482.72 | [446.444, 521.115] | 353.363 | [342.169, 364.233] |
| tau.4 | 353.412 | [333.737, 372.59] | 259.537 | [248.238, 270.149] |
| tau.5 | 430.821 | [403.804, 459.002] | 182.106 | [174.342, 189.982] |
| tau.6 | 243.005 | [214.896, 271.659] | 308.051 | [295.226, 316.926] |
| tau.7 | 378.525 | [341.233, 446.348] | 206.454 | [195.921, 219.556] |
| tau.8 | 211.58 | [142.955, 249.64] | 346.626 | [334.93, 356.425] |
| tau.9 | 641.962 | [559.345, 689.229] | 141.479 | [133.361, 146.888] |
| tau.10 | NA | NA | 234.009 | [224.291, 248.884] |
| tau.11 | NA | NA | 200.392 | [193.409, 207.805] |
| tau.12 | NA | NA | 568.935 | [558.553, 577.11] |
| tau.13 | NA | NA | NA | NA |
| mu.tau | 375.057 | [285.636, 473.536] | 275.794 | [190.848, 365.31] |
| sigma.tau | 144.288 | [95.171, 195.317] | 132.09 | [87.542, 192.112] |
| rho | 0 | [0, 0] | 0.092 | [0.081, 0.099] |
| delta | 53.435 | [26.502, 87.574] | 31.774 | [25.385, 44.667] |
| z | 4.626 | [1.219, 15.057] | 4.415 | [3.94, 5.029] |
| n | 626.953 | [167.313, 966.437] | 30.534 | [30.039, 30.963] |

Table S2 cont.

| param | Estimate_cvn_h1 | CrI_cvn_h1 | Estimate_cvn_h3 | CrI_cvn_h3 | Estimate_cvn_a | CrI_cvn_a |
|---|---|---|---|---|---|---|
| beta | 0.168 | [0.16, 0.173] | 0.183 | [0.175, 0.188] | 0.191 | [0.186, 0.192] |
| a | 0.833 | [0.719, 0.965] | 0.256 | [0.185, 0.341] | 0.247 | [0.175, 0.328] |
| gamma | 1838.565 | [1009.857, 2164.404] | 1203.187 | [451.036, 2083.336] | 2115.092 | [1110.749, 2186.642] |
| phi.1 | 149.082 | [144.465, 153.742] | 335.585 | [297.965, 366.861] | 180.771 | [162.07, 208.15] |
| tau.1 | 642.602 | [629.613, 654.437] | 177.286 | [146.827, 218.336] | 325.759 | [271.976, 348.863] |
| tau.2 | 205.911 | [192.8, 218.83] | 523.35 | [500.16, 540.947] | 372.167 | [353.686, 409.111] |
| tau.3 | 294.439 | [285.793, 300.702] | 159.663 | [146.027, 179.653] | 155.125 | [133.9, 172.873] |
| tau.4 | 524.594 | [518.301, 530.05] | 435.299 | [413.135, 456.222] | 269.896 | [253.35, 279.167] |
| tau.5 | 205.413 | [171.084, 219.182] | 522.005 | [480.89, 561.873] | 318.203 | [297.283, 334.928] |
| tau.6 | 641.763 | [621.899, 677.887] | 162.15 | [128.344, 202.17] | 213.084 | [196.382, 236.359] |
| tau.7 | 236.673 | [214.999, 254.437] | 338.527 | [295.489, 388.994] | 264.142 | [222.67, 276.276] |
| tau.8 | NA | NA | 351.874 | [300.288, 400.905] | 225.501 | [214.429, 257.518] |
| tau.9 | NA | NA | 441.123 | [403.96, 488.685] | 316.336 | [298.079, 368.323] |
| tau.10 | NA | NA | NA | NA | 371.921 | [324.457, 389.763] |
| tau.11 | NA | NA | NA | NA | NA | NA |
| tau.12 | NA | NA | NA | NA | NA | NA |
| tau.13 | NA | NA | NA | NA | NA | NA |
| mu.tau | 389.451 | [255.45, 516.839] | 342.122 | [241.176, 448.347] | 281.612 | [228.779, 336.961] |
| sigma.tau | 171.442 | [124.171, 198.271] | 157.211 | [102.039, 196.255] | 78.405 | [51.194, 125.008] |
| rho | 0.001 | [0, 0.002] | 0.038 | [0.02, 0.054] | 0.04 | [0.028, 0.069] |
| delta | 124.958 | [116.137, 137.217] | 110.023 | [72.501, 144.649] | 87.872 | [66.81, 134.304] |
| z | 56.98 | [15.589, 140.813] | 15.278 | [7.081, 24.767] | 5.79 | [2.677, 8.774] |
| n | 37.53 | [37.029, 37.952] | 66.517 | [66.046, 66.947] | 74.51 | [74.029, 74.956] |

Table S2 cont.

| param | Estimate_cvn_b | CrI_cvn_b | Estimate_cvn_ili+ | CrI_cvn_ili+ |
|---|---|---|---|---|
| beta | 0.205 | [0.203, 0.207] | 0.192 | [0.19, 0.196] |
| a | 0.185 | [0.122, 0.353] | 0.323 | [0.167, 0.389] |
| gamma | 481.412 | [425.981, 545.644] | 1928.155 | [1142.66, 2151.924] |
| phi.1 | 8.398 | [1.521, 23.074] | 177.39 | [166.168, 203.096] |
| tau.1 | 349.465 | [324.275, 372.821] | 193.317 | [159.741, 204.199] |
| tau.2 | 351.76 | [321.568, 378.151] | 163.56 | [150.759, 182.677] |
| tau.3 | 454.567 | [397.111, 513.8] | 261.387 | [243.654, 277.243] |
| tau.4 | 346.779 | [295.635, 397.551] | 426.726 | [411.332, 441.948] |
| tau.5 | 220.008 | [201.96, 242.985] | 104.897 | [92.831, 114.792] |
| tau.6 | 364.65 | [313.744, 428.312] | 433.923 | [424.594, 444.708] |
| tau.7 | 308.616 | [236.69, 367.031] | 101.936 | [88.342, 108.474] |
| tau.8 | 459.405 | [421.131, 503.683] | 62.77 | [25.945, 216.063] |
| tau.9 | 270.987 | [230.864, 305.734] | 242.213 | [196.819, 280.801] |
| tau.10 | NA | NA | 280.825 | [154.408, 321.02] |
| tau.11 | NA | NA | 199.535 | [163.814, 222.999] |
| tau.12 | NA | NA | 247.497 | [236.287, 276.292] |
| tau.13 | NA | NA | 433.547 | [347.872, 481.869] |
| mu.tau | 346.407 | [291.931, 401.659] | 245.652 | [158.136, 330.911] |
| sigma.tau | 83.061 | [59.233, 107.065] | 132.678 | [92.602, 189.25] |
| rho | 0 | [0, 0] | 0.075 | [0.022, 0.097] |
| delta | 84.246 | [40.348, 124.141] | 21.925 | [20.087, 41.58] |
| z | 49.152 | [22.444, 97.291] | 5.479 | [4.357, 13.579] |
| n | 666.257 | [608.921, 707.963] | 36.492 | [36.01, 36.962] |

Table S2 cont.

| param | Estimate_svn_h1 | CrI_svn_h1 | Estimate_svn_h3 | CrI_svn_h3 | Estimate_svn_a | CrI_svn_a |
|---|---|---|---|---|---|---|
| beta | 0.147 | [0.14, 0.159] | 0.164 | [0.157, 0.169] | 0.174 | [0.172, 0.176] |
| a | 0.779 | [0.638, 0.886] | 0.511 | [0.435, 0.612] | 0.394 | [0.365, 0.425] |
| gamma | 1101.764 | [433.149, 1783.258] | 1506.666 | [441.467, 2171.48] | 996.951 | [422.663, 1827.601] |
| phi.1 | 110.069 | [107.138, 113.098] | 283.145 | [276.217, 287.765] | 141.317 | [133.083, 143.726] |
| tau.1 | 190.05 | [171.514, 204.818] | 201.128 | [193.137, 209.227] | 370.163 | [364.032, 380.137] |
| tau.2 | 558.824 | [543.697, 577.895] | 440.132 | [429.685, 450.361] | 357.539 | [349.055, 368.017] |
| tau.3 | 434.418 | [427.103, 442.69] | 241.058 | [231.572, 251.02] | 320.812 | [310.713, 330.186] |
| tau.4 | 501.74 | [493.44, 510.24] | 445.803 | [437.655, 453.213] | 132.25 | [125.668, 137.961] |
| tau.5 | 255.689 | [249.182, 262.056] | 367.405 | [353.2, 379.515] | 292.813 | [286.72, 299.001] |
| tau.6 | 586.779 | [579.557, 595.028] | 298.909 | [287.484, 313.276] | 185.161 | [176.744, 206.752] |
| tau.7 | 304.001 | [286.879, 318.878] | 370.784 | [354.798, 382.459] | 259.986 | [240.005, 271.481] |
| tau.8 | 352.042 | [337.862, 369.197] | 309.471 | [297.523, 325.042] | 257.769 | [250.083, 266.139] |
| tau.9 | NA | NA | 430.514 | [423.145, 438.051] | 346.05 | [342.153, 351.473] |
| tau.10 | NA | NA | NA | NA | 318.549 | [313.296, 324.38] |
| tau.11 | NA | NA | NA | NA | 342.003 | [338.757, 345.314] |
| tau.12 | NA | NA | NA | NA | NA | NA |
| tau.13 | NA | NA | NA | NA | NA | NA |
| mu.tau | 396.38 | [285.792, 499.121] | 343.622 | [264.596, 416.428] | 288.314 | [228.975, 341.688] |
| sigma.tau | 149.467 | [94.359, 194.823] | 104.136 | [62.722, 171.826] | 85.1 | [53.091, 149.655] |
| rho | 0.007 | [0.003, 0.014] | 0.011 | [0.006, 0.016] | 0.083 | [0.072, 0.094] |
| delta | 157.216 | [142.94, 170.557] | 171.517 | [158.2, 179.284] | 82.34 | [77.524, 88.372] |
| z | 7.135 | [2.394, 20.677] | 12.363 | [8.707, 19.626] | 5.459 | [4.738, 6.286] |
| n | 44.505 | [44.03, 44.969] | 44.453 | [44.028, 44.946] | 38.519 | [38.039, 38.96] |

Table S2 cont.

| param | Estimate_svn_b | CrI_svn_b | Estimate_svn_ili+ | CrI_svn_ili+ |
|---|---|---|---|---|
| beta | 0.189 | [0.186, 0.192] | 0.196 | [0.196, 0.196] |
| a | 0.237 | [0.191, 0.306] | 0.177 | [0.142, 0.222] |
| gamma | 1072.865 | [379.967, 1432.675] | 1383.984 | [1257.423, 1515.013] |
| phi.1 | 223.612 | [205.174, 241.45] | 123.324 | [112.534, 132.443] |
| tau.1 | 393.507 | [367.261, 427.095] | 176.511 | [161.02, 190.601] |
| tau.2 | 344.179 | [309.728, 379.191] | 225.922 | [214.885, 236.952] |
| tau.3 | 431.73 | [379.443, 485.054] | 333.906 | [321.299, 346.066] |
| tau.4 | 263.112 | [209.569, 311.966] | 314.447 | [298.785, 330.366] |
| tau.5 | 409.787 | [392.065, 433.611] | 118.799 | [102.044, 133.588] |
| tau.6 | 319.227 | [296.331, 336.64] | 334.265 | [321.467, 345.511] |
| tau.7 | 406.191 | [387.526, 430.574] | 428.703 | [407.05, 445.735] |
| tau.8 | 311.58 | [286.729, 330.164] | 244.285 | [227.861, 264.812] |
| tau.9 | 432.722 | [395.368, 479.144] | 147.99 | [134.556, 161.591] |
| tau.10 | NA | NA | 192.001 | [179.336, 204.523] |
| tau.11 | NA | NA | 181.865 | [158.86, 202.553] |
| tau.12 | NA | NA | 162.568 | [148.897, 177.243] |
| tau.13 | NA | NA | 332.754 | [325.121, 341.56] |
| mu.tau | 368.233 | [318.662, 415.855] | 248.825 | [164.231, 302.453] |
| sigma.tau | 69.995 | [38.913, 130.288] | 109.64 | [72.81, 170.775] |
| rho | 0.001 | [0.001, 0.003] | 0.09 | [0.088, 0.096] |
| delta | 172.607 | [146.283, 179.469] | 53.382 | [41.965, 65.619] |
| z | 1.413 | [1.011, 2.97] | 4.315 | [3.946, 4.617] |
| n | 152.945 | [127.281, 156.603] | 293.527 | [293.041, 293.956] |

Table S2 cont.

| param | Estimate_nl_ili+ | CrI_nl_ili+ | Estimate_dk_ili+ | CrI_dk_ili+ |
|---|---|---|---|---|
| beta | 0.606 | [0.603, 0.609] | 0.592 | [0.589, 0.595] |
| a | 0.104 | [0.101, 0.107] | 0.14 | [0.138, 0.141] |
| gamma | 1108.097 | [1104.814, 1112.198] | 819.913 | [817.548, 822.245] |
| phi.1 | 378.017 | [376.747, 379.309] | 68.361 | [67.47, 69.33] |
| tau.1 | 419.328 | [417.147, 421.59] | 443.634 | [441.774, 445.872] |
| tau.2 | 270.94 | [268.885, 272.863] | 294.09 | [291.019, 297.414] |
| tau.3 | 445.864 | [443.243, 448.989] | 350.765 | [348.521, 352.96] |
| tau.4 | 327.584 | [325.317, 329.417] | 387.986 | [386.426, 389.915] |
| tau.5 | 332.592 | [330.474, 334.498] | 312.467 | [311.611, 313.309] |
| tau.6 | 362.87 | [360.893, 365.054] | 325.56 | [325.025, 325.981] |
| tau.7 | 430.998 | [429.129, 432.844] | 439.19 | [437.376, 440.76] |
| tau.8 | 279.099 | [276.963, 281.306] | 387.66 | [385.742, 390.104] |
| tau.9 | 372.487 | [369.759, 375.027] | 252.216 | [251.259, 253.204] |
| tau.10 | NA | NA | NA | NA |
| tau.11 | NA | NA | NA | NA |
| tau.12 | NA | NA | NA | NA |
| tau.13 | NA | NA | NA | NA |
| mu.tau | 367.82 | [319.517, 437.313] | 361.868 | [354.338, 383.569] |
| sigma.tau | 73.984 | [44.537, 115.802] | 56.687 | [48.103, 63.534] |
| rho | 0 | [0, 0] | 0 | [0, 0] |
| delta | 81.213 | [77.874, 84.72] | 140.391 | [138.137, 142.53] |
| z | 21.411 | [2.441, 38.193] | 4.887 | [1.3, 8.082] |
| n | 93.494 | [93.037, 93.957] | 51.559 | [51.029, 51.961] |

Table S2 cont.

| param | Estimate_us1_ili | CrI_us1_ili | Estimate_us9_ili | CrI_us9_ili |
|---|---|---|---|---|
| beta | 0.522 | [0.517, 0.529] | 0.462 | [0.451, 0.473] |
| a | 0.204 | [0.19, 0.218] | 0.061 | [0.059, 0.063] |
| gamma | 2010.36 | [1998.572, 2025.669] | 1980.025 | [1939.14, 2020.319] |
| phi.1 | 31.898 | [30.216, 33.383] | 1.628 | [1.033, 5.465] |
| tau.1 | 316.194 | [313.982, 318.249] | 301.611 | [297.163, 304.792] |
| tau.2 | 392.1 | [389.649, 394.414] | 399.238 | [395.635, 402.68] |
| tau.3 | 278.126 | [274.728, 281.757] | 334.493 | [330.899, 337.669] |
| tau.4 | 358.321 | [354.473, 362.173] | 350.476 | [345.204, 355.787] |
| tau.5 | 356.427 | [350.736, 361.929] | 363.935 | [360.939, 366.945] |
| tau.6 | 423.876 | [418.016, 429.741] | 387.503 | [382.776, 392.137] |
| tau.7 | 367.077 | [365.354, 368.868] | 360.219 | [356.885, 364.072] |
| tau.8 | 360.142 | [357.41, 362.579] | 361.724 | [358.995, 364.287] |
| tau.9 | 378.462 | [374.209, 382.694] | 356.261 | [352.223, 360.035] |
| tau.10 | NA | NA | NA | NA |
| tau.11 | NA | NA | NA | NA |
| tau.12 | NA | NA | NA | NA |
| tau.13 | NA | NA | NA | NA |
| mu.tau | 361.443 | [330.276, 394.386] | 357.365 | [333.714, 381.871] |
| sigma.tau | 47.388 | [30.691, 81.453] | 32.826 | [20.275, 59.302] |
| rho | 0 | [0, 0] | 0 | [0, 0.001] |
| delta | 56.682 | [52.724, 61.197] | 179.704 | [178.521, 179.982] |
| z | 22.361 | [2.577, 44.932] | 3.674 | [1.033, 8.248] |
| n | 59.305 | [58.141, 59.952] | 52.696 | [30.676, 69.111] |
